# The New York Genome Center ALS Consortium resource integrates postmortem tissue transcriptomics and whole genome sequencing to empower biological discovery

**DOI:** 10.64898/2026.04.29.26350889

**Authors:** Jack Humphrey, Ali Oku, Marta Byrska-Bishop, Anna O. Basile, Uday S. Evani, André Corvelo, Alex Tokolyi, Kailash BP, Aline Réal, Yebin Kim, Marielle L. Bond, Wayne E. Clarke, Rui Fu, Heather Geiger, Sei Chang, Tatsuhiko Naito, Beomjin Jang, Rajeeva Musunuri, Winston H Dredge, Rashid Al-Abri, Benjamin N Hoover, Dina Manaa, Jaime McClintock, Faith P Singh, Maria H Pedersen, Alexi Runnels, Nadia Propp, Samantha Fennessey, Hong-Hee Won, Michael C. Zody, Giuseppe Narzisi, Nicolas Robine, Tuuli Lappalainen, Delphine Fagegaltier, Gamze Gürsoy, David A. Knowles, Towfique Raj, NYGC ALS Consortium, Matthew B Harms, Hemali Phatnani

## Abstract

Amyotrophic lateral sclerosis (ALS) is a devastating neurodegenerative disease with substantial genetic and clinical heterogeneity that impedes therapeutic development. Large-scale multi-tissue genomic resources have transformed the study of neuropsychiatric and neurodegenerative diseases, but no equivalent resource exists for ALS. Here we present the full NYGC ALS Consortium dataset, combining whole-genome sequencing from 4,746 donors and bulk RNA-seq from 2,574 samples across 8 brain and spinal cord regions from 695 donors across the ALS disease spectrum. Our catalogue of small variants, structural variants, and short tandem repeats identified likely pathogenic mutations in 15.6% of ALS cases. Gene expression and mRNA splicing analysis across 5 major tissues reveals shared and region-specific features, highlighting microglial and T-cell dysregulation in the spinal cord. Mapping the genetic regulation of expression and splicing across tissues identified associations with 6 ALS risk loci, whereas allele-specific rare variant analysis detected expression effects for *C9orf72* and *OPTN*. All data are immediately publicly available.

## Introduction

Amyotrophic lateral sclerosis (ALS) is a devastating neurodegenerative disease, classically characterized by progressive loss of motor neurons in the motor cortex and spinal cord, resulting in total paralysis and death. In the past decades, there has been substantial progress in identifying both common genetic risk factors and causal mutations for ALS^1–3^, with genetic variation acting across different cell-types and brain regions, beyond motor neurons^4–6^. Additionally, ALS patients have substantial clinical heterogeneity, which may represent distinct molecular subtypes^7–9^. This represents a significant roadblock for the development of new therapeutics. Addressing this requires integrating genetic data with transcriptomic data across the multiple tissues affected in ALS, at a scale sufficient to map the regulatory effects of common and rare genetic variants.

Advances in sequencing technologies and data integration methods have enabled genomic approaches to identify disease mechanisms. Pioneering integrative genetic and bulk transcriptomic studies from the GTEx consortium enabled the large-scale study of the genetic regulation of gene expression and mRNA splicing in postmortem human tissues^10^. This approach has been successfully applied to brain tissues across development^11^, neuropsychiatric disease^12–14^, and particularly in Alzheimer’s disease^15–17^, where further integration with clinical and neuropathology data is enabling molecular subtyping and disease trajectories across brain regions^18,19^. ALS, being a comparatively rare disease, has lagged behind these efforts. The New York Genome Center (NYGC) ALS Consortium was formed in 2015 to address this challenge, bringing together clinicians, basic scientists, geneticists, and computational biologists globally from 41 institutions. This partnership established a framework to generate whole genome sequencing (WGS) and bulk tissue RNA sequencing (RNA-seq) data and apply functional genomics approaches to survey the genetic, molecular, and cellular basis of ALS. We have built a comprehensive and openly accessible resource of genomic and clinical data from ALS patients to enable the research community to answer critical scientific questions, accelerate biological discovery, and advance the development of novel therapeutics. We have continuously made data available over the construction of the cohort, enabling multiple independent groups to generate findings in advance of this publication^2,3,7–9,20–28^, demonstrating the consortium’s value to the field.

We previously published a focussed study of the ALS spinal cord using 380 RNA-seq samples from 280 donors with paired WGS^6^. Here we present the full NYGC ALS Consortium dataset, comprising WGS on 4,746 donors and RNA-seq from 8 major brain and spinal cord tissues from a cohort of 695 individuals presenting with ALS, other neurological diseases, and non-neurological controls. Maximising the utility of WGS, we comprehensively survey rare and common variation across small variants, structural variants, and short tandem repeats, identifying a genetic mutation for 15.6% of ALS patients. Assembling the largest set of ALS tissue transcriptomics to date, we assess shared and tissue-specific changes in gene expression and mRNA splicing in ALS patients, and use paired clinical information to identify clinical and genetic subtype associations, revealing microglia and T-cell dysregulation in the spinal cord. We augment our bulk tissue insights with tissue-matched single-cell data to predict cell-type composition changes, identifying the cerebellum as a clear regional outlier in transcriptomic response to disease. Finally, we integrate the two data types using quantitative trait locus (QTL) mapping and allele-specific expression (ASE) approaches to demonstrate how common risk variants, short tandem repeats, and rare ALS mutations can affect the transcriptome, validating *C9orf72* repeat expansion effects on *C9orf72* gene expression and splicing with three orthogonal approaches. Collectively, our findings establish the NYGC ALS Consortium cohort as a powerful resource for integrating genetic variation with cellular and clinical features of ALS.

## Results

### The NYGC ALS Consortium cohort

Blood and autopsy tissue samples were obtained from 41 ALS clinics and research institutions spanning the USA, the UK, the Netherlands, Greece, and Israel (**Fig. 1A)**. All samples were sent for library preparation and sequencing at the NYGC. No selection criteria was imposed to maximise potential sample size. Clinical data on each patient as provided by each institution was harmonized by a team at NYGC. Ultimately, site of symptom onset (limb, bulbar, other), age at symptom onset, and age at death were available for the majority of donors. WGS was performed on 4,828 donors, with a final set of 4,746 passing quality control. Bulk tissue RNA-seq was performed on 2,574 samples originating from 695 unique donors with matching WGS and clinical records. The cohort consists primarily of 3,293 donors with classical/typical ALS and 443 non-neurological controls (**Fig. 1B; Table S1**) with 191 donors with concurrent ALS and frontotemporal dementia (ALS-FTD), 146 donors with pure FTD, 251 donors with other motor neuron diseases, including primary lateral sclerosis and spinal bulbar muscular atrophy, and 422 donors with other neurological diseases. Genetic ancestry estimation and principal component analysis demonstrated that the cohort is predominantly (89.7%) of European descent, with 285 donors having a mixture of ancestries (**Fig. 1C)**. RNA-seq data was generated from 5 major regions of the brain and 3 spinal cord sections, a primary site of ALS pathology, with each donor contributing a median of 3 tissues (**Fig. 1D**). Principal component analysis of the expression data for the eight largest tissues revealed three major groups (**Fig. 1E**).

**Figure 1.**
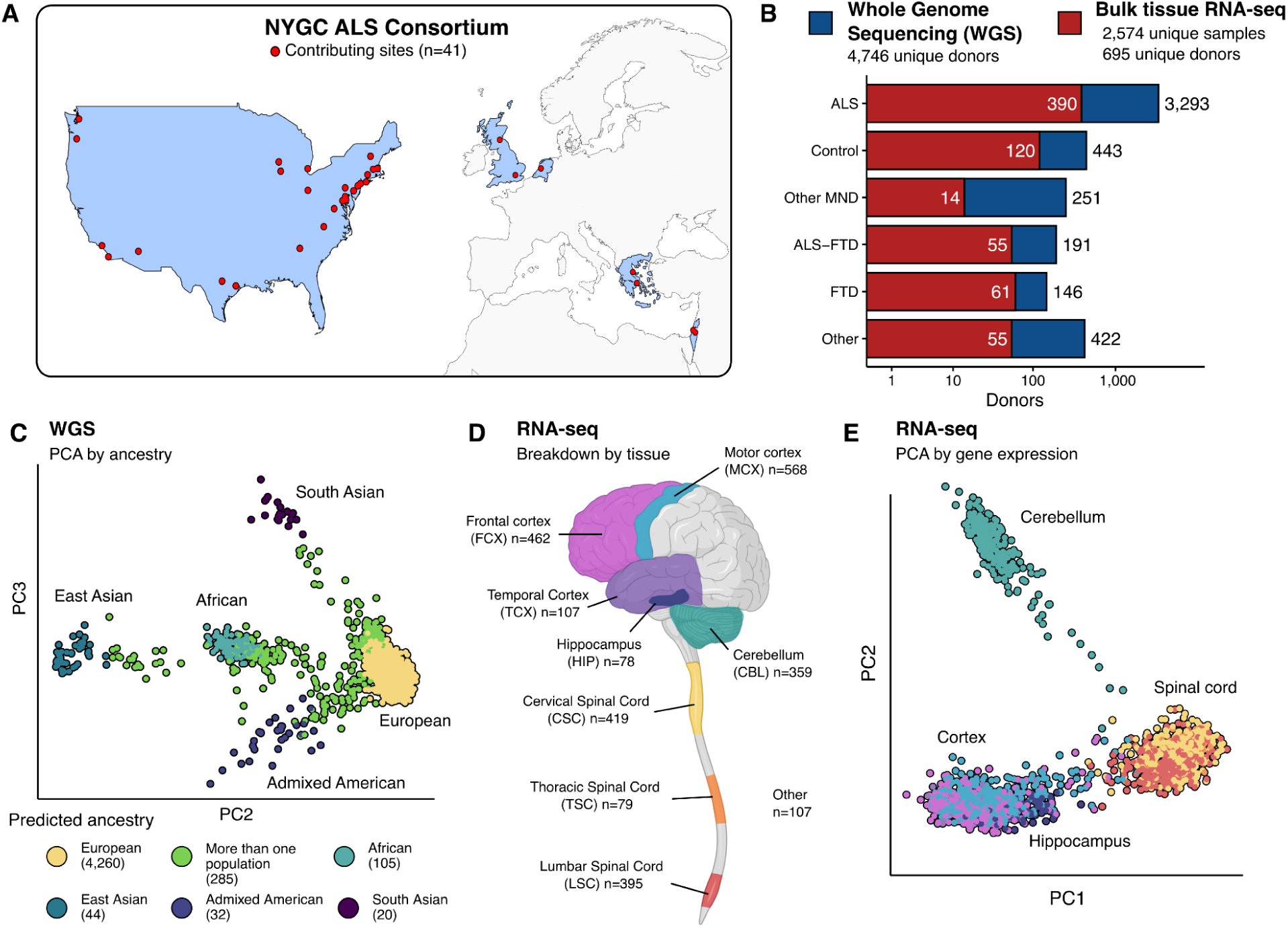
| The New York Genome Center ALS Consortium. **A)** Blood and tissue samples were contributed by 41 sites across 5 countries. **B)** Overview of diagnostic groups in the WGS and RNA-seq. **C)** Genetic principal component analysis (PCA) of the WGS cohort when projected on the 1000 Genomes superpopulations to estimate ancestry. **D)** Breakdown of brain and spinal cord tissues used for bulk RNA-seq. **E)** Gene expression PCA of the top 8 tissues shows clear separation by major region.

### Small variant discovery in WGS data

We performed WGS on 4,828 samples using either PCR+ or PCR-free library preparation kits and across three Illumina sequencing platforms (**Fig. 1B; Table S1; Methods Table 1**). 82 samples failed sample QC and were dropped from further analysis, resulting in 4,746 WGS samples that went into variant calling. Across all samples, we discovered and jointly genotyped 121,031,405 small variants passing variant quality score recalibration (VQSR PASS) using GATK v3.5^29,30^, including 105,266,396 single nucleotide variants (SNV) and 15,765,009 short insertions and deletions (INDELs) spanning the allele frequency spectrum (**Fig. 2A**). Approximately 11.5% (n=12,139,020) of discovered SNVs and 8.6% of INDELs (n=1,352,473) were novel with respect to dbSNP b156 (**Fig. 2A**). On average, we called 3.763-4.548 million SNVs per genome across the 6 genetically-inferred ancestry groups. Sample-level SNV count distributions followed the expected pattern of genetic diversity, with samples in the African (AFR) ancestry group carrying the highest number of variants (**Fig. S1A**). Per-genome INDEL counts depended on the type of library preparation kit used during sample processing, with samples processed using PCR-based kits carrying a considerably higher number of INDELs compared to PCR-free samples. Specifically, among European samples, we called an average of 0.942 million INDELs in PCR-free samples, 1.040 million INDELs in KAPA Hyper PCR+ samples, and 1.184 million INDELs in Nano PCR+ samples (**Fig. S1B**). As expected, the AFR samples carried the highest number of INDELs among ancestry groups (1.13-1.34M), depending on the library preparation type. This effect was largely resolved by applying stringent quality control (QC) criteria, defined based on VQSR, genotype call rate, Hardy-Weinberg Equilibrium (HWE), and masking of low-complexity regions (LCR) of the genome to all sites in the call set (**Figure S1B**). At the cohort-level, 101,076,094 (83.5%) small variants, 95,907,371 (91.1%) SNVs and 5,168,723 (32.8%) INDELs passed our stringent QC criteria and were used for downstream analysis (**Figure 2A; Methods Table 2**). The high fraction of INDELs excluded by stringent QC primarily match sites within difficult to sequence low-complexity regions of the genome^31^. We then leveraged 11 mother-father-child trios in the WGS cohort to assess the quality of called genotypes by computing mendelian error rate (MER) for each variant type. The average MER was 1.54% for rare (minor allele frequency (MAF) < 1%) and 0.77% for common (MAF ≥ 1%) SNVs, indicating high quality of VQSR passing genotypes (**Figure S1C**). Applying stringent QC criteria improved MER by 3.8-fold for rare SNVs and 7-fold for common variants, decreasing to 0.41% and 0.11%, respectively. In the case of INDELs, the average MER improved over 30-fold (decreased from 18.88% to 0.62%) for rare and over 44-fold (decreasing from 7.99% to 0.18%) for common INDELs. This improvement was primarily due to LCR filtering, as the intermediate filtering (all stringent QC criteria except for LCR filtering) condition did not show as significant of an improvement in MER (**Figure S1C**).

**Methods Table 1.**
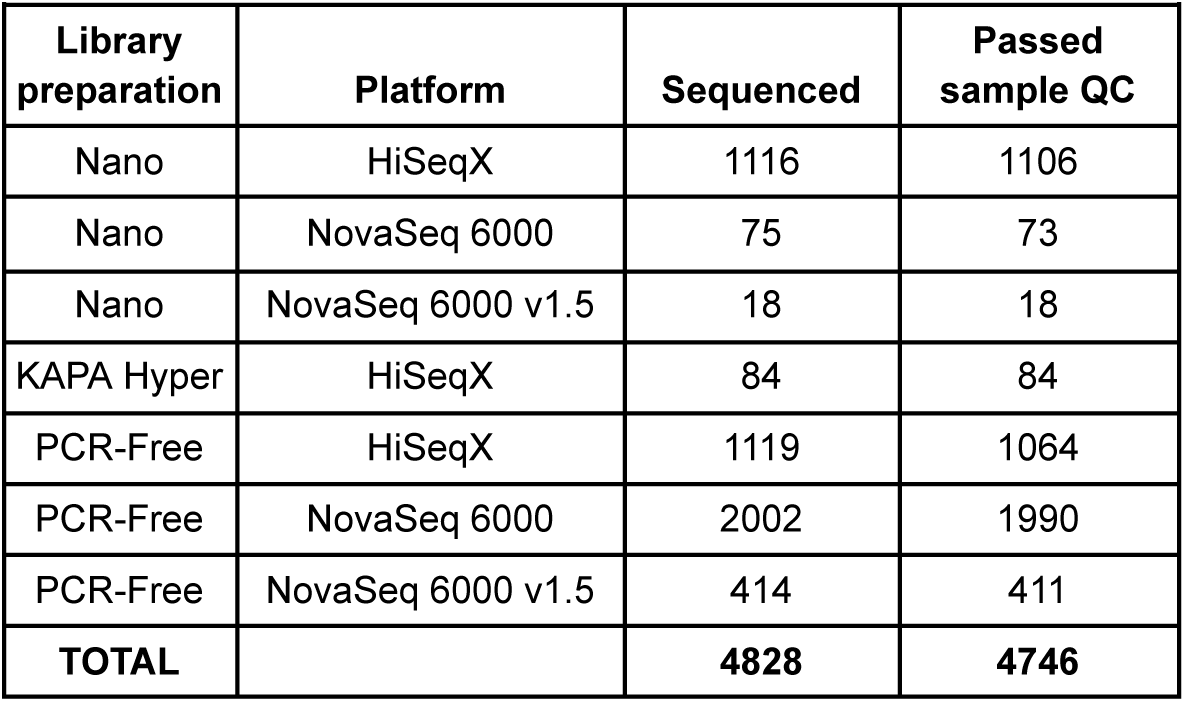
WGS library prep kits and platforms.

| **Methods Table 1. WGS library prep kits and platforms** |  |  |  |
| --- | --- | --- | --- |
| **Library preparation** | **Platform** | **Sequenced** | **Passed sample QC** |
| Nano | HiSeqX | 1116 | 1106 |
| Nano | NovaSeq 6000 | 75 | 73 |
| Nano | NovaSeq 6000 v1.5 | 18 | 18 |
| KAPA Hyper | HiSeqX | 84 | 84 |
| PCR-Free | HiSeqX | 1119 | 1064 |
| PCR-Free | NovaSeq 6000 | 2002 | 1990 |
| PCR-Free | NovaSeq 6000 v1.5 | 414 | 411 |
| **TOTAL** |  | **4828** | **4746** |

**Figure 2.**
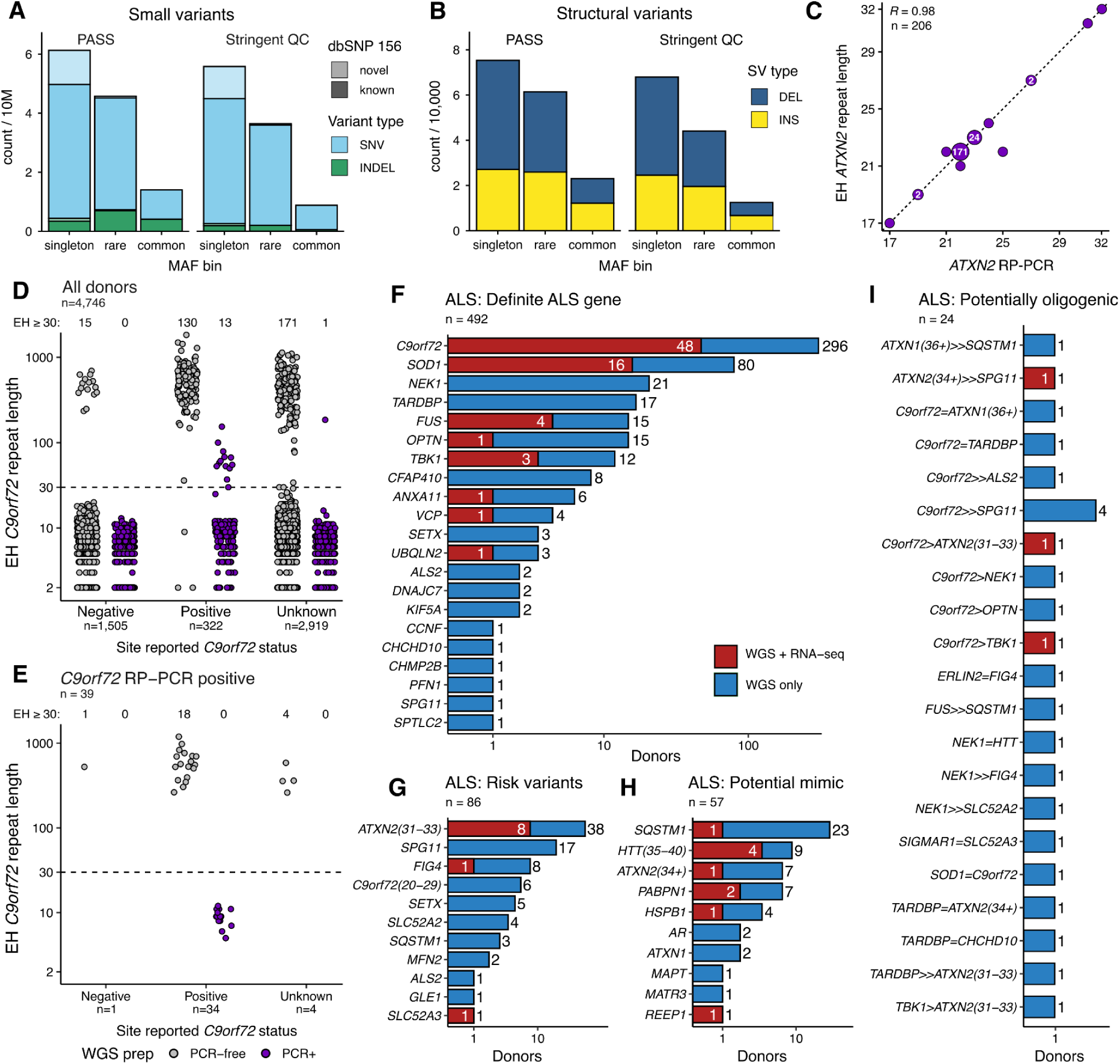
| Variant discovery and ALS mutation annotation across 4,746 WGS samples. **A)** Cohort-level counts of small variants stratified by MAF bin, filtering condition (VQSR-passing or stringent QC), variant type (SNV & INDEL), and comparison against dbSNP build 156. Singleton: minor allele count (MAC) = 1, rare: MAF < 1%, common: MAF ≥ 1%, **B)** Cohort-level counts of structural variants (SVs) stratified by MAF bins and filtering condition. **C**) Correlation between ExpansionHunter-estimated repeat length and RP-PCR estimated repeat length for *ATXN2* for 206 alleles in 103 samples. **D)** ExpansionHunter-estimated repeat length distributions for *C9orf72* in 4,746 samples, split by site-reported *C9orf72* expansion status and WGS library preparation. **E**) Estimated *C9orf72* repeat lengths for *C9orf72* RP-PCR positive samples. **F-I)** Summary of mutations discovered in 659 ALS patients. **F)** Variants in definite ALS genes. **G)** Risk variants, including intermediate length expansions and single copies of known recessive genes. **H)** Potential ALS mimics. **I)** Potentially oligogenic donors. Driver variant in pair denoted by >. = refers to both genes being equally causal. DEL: large deletions, INS: large insertions. EH: ExpansionHunter.

### Structural variant discovery

We generated an ensemble SV call set across the 4,746 cohort samples. For each sample, large insertions and deletions were discovered using Manta^32^ and Absinthe^33^, genotyping each set across the cohort with Paragraph^34^. We combined these callsets with mobile element insertions called by MELT^35^ and insertion and deletion calls from the HGSVC project^36^, genotyped with both Paragraph and Pangenie^37^ to produce the final call set of 159,718 variant loci, composed of 65,237 insertions and 94,481 deletions (**Fig. 2B; Methods Table 2**). On average, 8,805 fully cohort-genotyped SVs were called in each genome (4,926 insertions and 3,879 deletions), with the greatest number of SVs per genome observed in individuals of predominantly African ancestry (**Fig. S2A**). These numbers are comparable to high coverage short-read sequencing results from the 1000 Genomes Project^38^. Similarly to short variants, most SVs were either singletons or rare, with approximately 18.68% of the insertions and 11.47% of the deletions considered common (MAF >= 0.01) (**Fig. 2B, Fig. S2B**). 49.29% of the SVs overlapped RefSeq-curated genes, including promoter regions. Of the 5.24% overlapping exons, almost half (47.26%) overlap coding sequences (**Fig. S2C**). In terms of SV size, signatures were observed for ALU (∼300 bp), SVA (∼1.2 kb) and LINE1 (∼6 kb) insertions as well as ALU deletions (**Fig. S2D**). Of the most common high impact SVs, as defined by Variant Effect Predictor (VEP)^39^, unsurprisingly, were deletions responsible for transcript ablation, most prominently splice sites, and insertions responsible for feature elongation (**Fig. S2E**). Similarly to small variant evaluation, we used MER calculation across 11 trios in the call set to assess the quality of SV genotypes. SVs displayed low MER overall, in line with that observed for SNVs, indicating high quality of called genotypes. Specifically, MER among rare SVs ranged from 1.84% to 2.23%, on average, and was similar across the 3 filtering conditions (**Fig. S2F**). Common SVs showed improvement in MER upon stringent QC filtering, decreasing nearly 2-fold, from 1.12% to 0.57%.

**Methods Table 2.** Total variant discovery.

| **Methods Table 2. Total variant discovery** |  |  |
| --- | --- | --- |
| **Variant type** | **PASS** | **Stringent QC** |
| **Total small variants** | **121,031,405** | **101,076,094** |
| SNV | 105,266,396 | 95,907,371 |
| INDEL | 15,765,009 | 5,168,723 |
| **Total SVs** | **159,718** | **124,475** |
| DEL | 94,481 | 73,575 |
| INS | 65,237 | 50,900 |

**Methods Table 3.** Models used for differential expression in each tissue.

|  |  |
| --- | --- |
| **Tissue** | **Covariates used in design** |
| CBL | Library prep method, mean read length, specimen collection site |
| MCX | Library prep method, mean read length, percent coding bases |
| TSC | Library prep method, median 5' bias, percent chimeras |
| LSC | Specimen collection site, prep, percent chimeras |
| CSC | Library prep method, mean read length, specimen collection site |
| FCX | Library prep method, mean read length, percent chimeras |
| TCX | Library prep method, percent chimeras, median 5' bias |
| HIP | Median 5' bias, median 3' bias, percent coding bases |

### Short tandem repeat discovery

Multiple ALS-associated genes harbor pathogenic STR expansions, including *C9orf72* and *ATXN2*. Accurate estimation requires PCR-free WGS. Using ExpansionHunter v5^40^ (EH), we analyzed 4,746 cohort samples across 68 clinically implicated STR loci and summarized repeat length distributions and pathogenic expansion frequencies (**Figure S3A**). To assess accuracy relative to the gold standard repeat-primed PCR (RP-PCR), we compared repeat length estimates from EH for *ATXN2* and *C9orf72* in samples with available RP-PCR data. ATXN2 repeat lengths estimated by EH showed strong agreement per allele with RP-PCR (n=206 alleles; Pearson’s correlation coefficient = 0.98, **Fig. 2C**). EH identified 329 donors with a *C9orf72* expansion (≥30 repeats^41^). While this included both PCR-free (n=316) and PCR-based (n=13) donors, EH cannot accurately estimate *C9orf72* repeat lengths in PCR-based WGS. While 322 donors were reported by their submitting site as being positive for a *C9orf72* repeat expansion, we could confirm this with EH in 142 donors (130 PCR-free; 12 PCR-based). We noted some discrepancies, as 15 site-reported negative donors were EH positive, while 3 site-reported positive donors with PCR-free WGS were EH negative (**Fig. 2D**). We used *C9orf72* RP-PCR to determine the presence of expanded alleles in 240 samples, confirming *C9orf72* expansion in 39 samples (23 PCR-free and 16 PCR-based library preparation samples). Only the PCR-free donors were both EH positive and RP-PCR confirmed (**Fig. 2E**). Acknowledging these limitations, we used a consensus of EH calls and RP-PCR validation, using positive site reports only in the PCR-based samples to annotate a final total of 506 donors as having *C9orf72* pathogenic expansions.

### Identifying known ALS mutations in 4,746 donors

To assess the functional impact of variants called across the 4,746 donors, we annotated the alleles with VEP^39^. Across all called SNVs and INDELs, we found a total of 721,908 missense, 412,436 synonymous, and 66,023 predicted loss-of-function mutations, including 16,890 stop-gain, 29,155 splice mutations and 19,978 frameshift mutations (**Figure S1D**). We then manually annotated mutations to a broad set of 154 genes associated with ALS, FTD, or reported mimic diseases. Definitive ALS genes were defined using a combination of the ClinGen Gene Curation Expert Panel (GCEP) list and other historical sources. The majority (n=492/659) of genetic ALS patients presented a single dominant mutation in one of 21 known ALS genes, with *C9orf72* expansions (≥30 repeats; n=296; 9% of all ALS) and *SOD1* missense mutations (n=80; 2.4%) being the most frequent (**Fig. 2F**). 86 ALS patients carried intermediate repeats in *ATXN2* (31-33; n=38) or *C9orf72* (20-29; n=6), or a single mutation in a known recessive ALS gene, including 17 patients heterozygous for *SPG11* mutations (**Fig. 2E**). *SPG11* is a known recessive gene for juvenile ALS^42^, but all heterozygous carriers had an age of onset greater than 45 years of age. A total of 57 donors diagnosed with ALS carried mutations associated with other neurological diseases, including 23 with mutations in *SQSTM1,* associated with Paget’s disease of bone^43^, and 9 carriers of repeat expansions in *HTT*, associated with Huntington’s disease (**Fig. 2D**). All 9 HTT expansion carriers have between 35 and 40 copies, which has been associated with reduced Huntington’s disease penetrance^44^. Finally, we identified 24 ALS patients carrying two ALS mutations, with 12 donors carrying a pathogenic *C9orf72* repeat expansion alongside another variant (**Fig. 2E**). In total, 516 ALS patients carried at least one definite ALS-associated mutation, for a total genetic annotation rate in ALS of 15.6% (516/3,293), with ALS patients with paired RNA-seq exhibiting a slightly higher annotation rate (20%, 78/390; *P* = 0.032, chi-squared test). We note that the additional 318 patients with annotated mutations include 62 ALS-FTD donors (32.4% annotation rate; 62/191), 66 FTD donors (45.2%; 66/146), and 46 non-ALS motor neuron disease donors (18.3%; 46/251). In line with previous studies, *C9orf72* repeat expansions were the predominant mutation in both ALS-FTD (n=53) and FTD (n=46; **Fig. S4**).

### Differential gene expression uncovers shared signal of ALS across tissues

Using the bulk tissue RNA-seq, we compared ALS donors to non-neurological controls for differential gene expression analysis. The 8 largest contributing tissues included both affected and less affected regions in ALS. They included the cerebellum (**CBL**; 206 cases, 39 controls), frontal cortex (**FCX**; 266 cases, 74 controls), motor cortex (**MCX**; 286 cases, 39 controls), temporal cortex (**TCX**; 32 cases, 27 controls), hippocampus (**HIP**; 52 cases, 14 controls), cervical spinal cord (**CSC**; 293 cases, 54 controls), lumbar spinal cord (**LSC**; 277 cases, 53 controls), and thoracic spinal cord (**TSC**; 48 cases, 9 controls). To maximize discovery power in the motor cortex we combined the three annotated regions (medial, lateral, and unspecified) into a single tissue-cohort, adjusting for repeated donors with a linear mixed model. In each tissue we identified technical sources of variation (**Fig. S5**), including library preparation batch, submitting site, RNA integrity number (RIN) and quality metrics derived directly from RNA-seq data, and included them as covariates in distinct models for each tissue (**Methods Table 2**). Gene expression changes affected primarily the cortical and spinal cord regions as expected, with some significant alterations also affecting the cerebellum. At a false discovery rate (FDR) threshold of 5%, the number of differentially expressed genes (DEGs) were as follows: MCX (8,759), CSC (9,211), LSC (9,449), FCX (10,403), CBL (3,134), HIP (19), TCX (2), and TSC (482) (**Fig. 3A; Table S4**). The discovery rate was strongly correlated with the effective sample size (**Fig. S6**). We removed HIP, TCX, and TSC from the remaining analyses due to low discovery power. Functional enrichment of DEGs was carried out using gene set enrichment analysis (GSEA) using the Gene Ontology (GO) functional databases. In the spinal cord, GO terms relating to immune signaling and inflammatory response were upregulated while pathways relating to neuronal development and cytoskeletal organization were downregulated. Cortical samples presented GO signatures consistent with heightened cellular metabolism and protein processing, while cell signaling and sensory perception pathways were decreased (**Table S5).**

**Figure 3.**
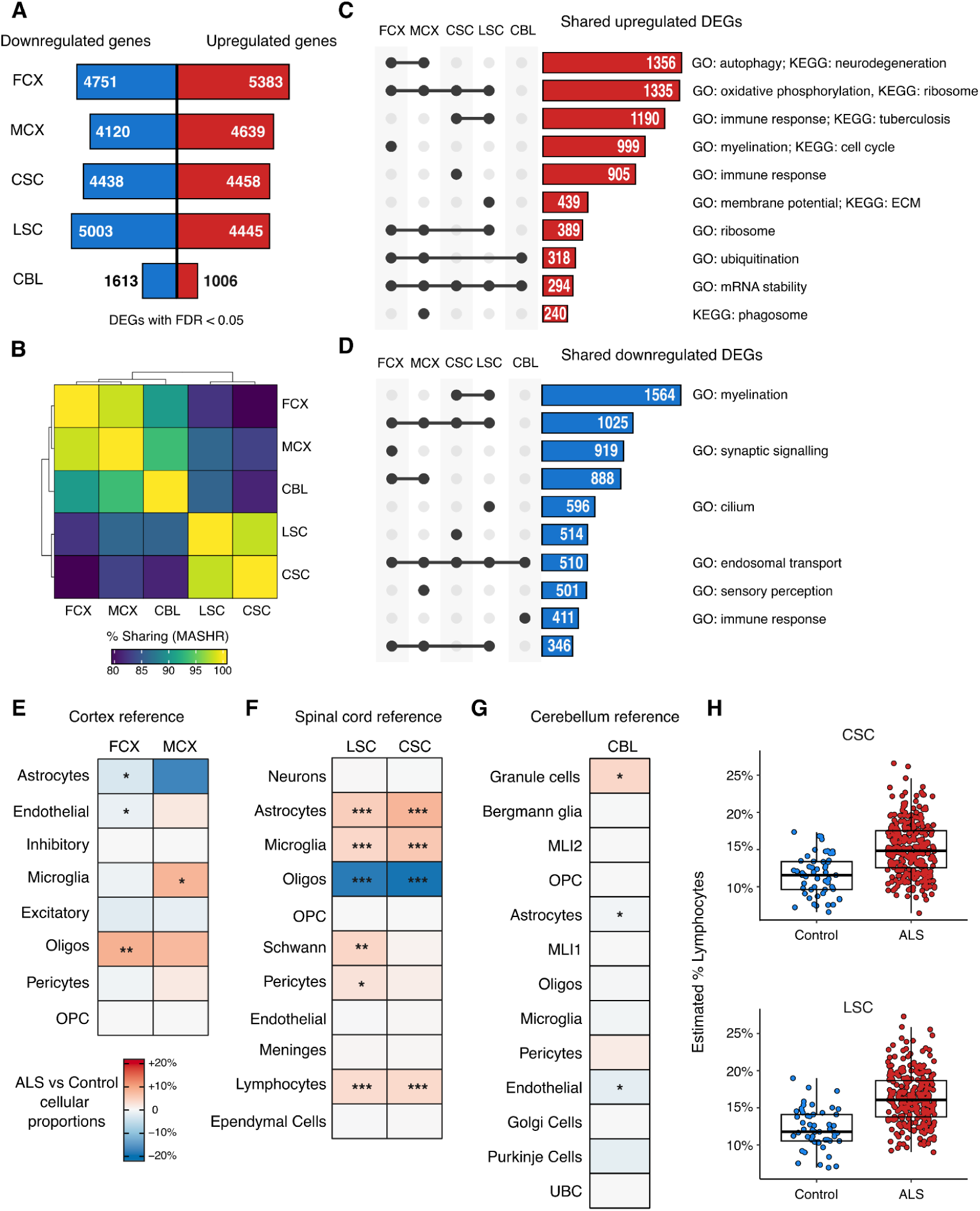
| Differential gene expression across tissues identifies pathway and cell composition effects. **A**) Numbers of differentially expressed genes (DEGs) split by direction of effect in each tissue at FDR < 0.05. **B**) Multi-adaptive shrinkage (MASH) estimates of shared DEGs across tissues. **C-D)** Modified upset plots intersecting upregulated (**C**) and downregulated (**D**) genes across tissues. Dots and lines represent specific intersections. Numbers refer to numbers of genes in each intersection. Gene sets manually annotated with GO and KEGG pathways. All reported pathways at Bonferroni-adjusted P < 0.05. Intersections without highlighted pathways had no significant enrichment. **E-G**) Differential cell composition as assessed by deconvolution using tissue-specific reference data from the frontal cortex^47^ (**E**), spinal cord^48^ (**F**), and cerebellum^49^ (**G**). P-values and effect sizes from linear regression. All P-values Bonferroni adjusted for the number of tested cell-type sets in each tissue reference. **H**) Lymphocyte expression is increased in ALS in both spinal cord regions. *** P < 0.001; ** P < 0.01; * P < 0.05.

We next sought to identify signatures across tissues. Using multivariate adaptive shrinkage^45^ on the five largest selected tissues, two main clusters emerged that reflect shared dysregulation: one cluster consisted of the CSC and LSC samples, while the FCX, MCX, and CBL formed the other (**Fig. 3B**). Next, we separated significant DEGs based on the direction of the fold changes to identify shared genes with consistent signals across tissues. We then performed over-representation analysis of each intersection using GO and KEGG^46^ to identify shared pathways (**Table S6**). We observed 1,356 upregulated genes shared uniquely between the FCX and MCX enriched in autophagy (GO) and neurodegeneration (KEGG) terms, 1,335 upregulated genes shared across the MCX, FCX, LSC, and CSC enriched in oxidative phosphorylation terms, and 1,190 upregulated genes shared between the CSC and LSC enriched in immune-related terms (**Fig. 3C**). Additionally, we identified 1,564 downregulated genes shared between the LSC and CSC enriched in myelination terms, 1,025 downregulated genes shared across the MCX, FCX, LSC, and CSC and 888 downregulated genes shared between the FCX and MCX (**Fig. 3D**). The shared downregulated genes were less frequently associated with specific GO or KEGG terms.

We hypothesized that a high proportion of the DEGs observed were due to cellular composition changes that occur during disease. We performed cell-type deconvolution using matching single-nucleus RNA-seq (snRNA-seq) reference data for each tissue type^47–49^ (**Fig. 3E-G, Table S7**). Deconvolution of spinal cord gene expression data revealed higher proportions of astrocytes, lymphocytes, and microglia, and a lower proportion of oligodendrocytes in ALS samples compared to controls (**Fig. 3E**). While we previously reported shifts in cell composition in the ALS spinal cord using a cortical snRNA-seq reference^6^, the use of a matched spinal cord snRNA-seq reference^48^ also identified an increase in lymphocytes (**Fig. 3H**). We note that infiltrating T-cells in the spinal cord has been reported in human patients^50^. In the FCX, ALS samples exhibited lower proportions of astrocytes and endothelial cells, and a higher proportion of oligodendrocytes compared to controls (**Fig. 3F, Table S8**). Finally, in the CBL, ALS samples exhibited a slightly higher proportion of neurons compared to controls (**Fig. 3G**). An orthogonal approach using GSEA with marker genes produced similar results (**Fig. S7, Table S9**). Therefore, while strong changes in cellular composition with accompanying changes in marker gene expression can be observed in the spinal cord, cell composition changes in the cortex and cerebellum are more subtle, likely owing to the increased complexity of cell subtypes in the brain.

### Gene expression associations with clinical and genetic heterogeneity

A key asset of the cohort is the harmonized clinical metadata on ALS patients made available through consortium-led efforts. To identify potential predictive signatures, we computed associations between gene expression and the following categorical or continuous clinical variables within the ALS patient tissues: 1) presence of *C9orf72* repeat, assayed from WGS; 2) site of symptom onset, characterized as either limb or bulbar; 3) age at first symptom onset; 4) age at death; 5) disease duration, taken as the difference between age at death and symptom onset.

We observed widely different numbers of DEGs associated with clinical variables and genetic subtypes (**Fig. 4A; Tables S10-15**). Comparing *C9orf72* repeat expansion carriers with non-carriers identified 1,236 DEGs in the CSC but fewer than 50 DEGs in every other tissue, with the *C9orf72* gene being downregulated in *C9orf72* carriers in 4/5 tissues (**Fig. 4B**). Across all tissues, few genes correlated with the site of symptom onset. In contrast, age-related DEGs were observed in the CBL, with 1,953 DEGs associated with age at death and 2,380 with age at onset, more than all other tissues combined for those categories. Disease duration DEGs were identified in all tissues but MCX, with the largest numbers seen in CSC (1,484) and CBL (345). We compared DEGs across all tissues by computing pairwise correlations of LFC values associated with age-related clinical factors. LFCs derived from disease duration formed a single cluster across all tissues, indicating a shared underlying signal. Within this cluster, the frontal and motor cortices grouped together, while the spinal cords formed a distinct subcluster, with the CBL clustering separately. In contrast, LFCs from other age-related clinical associations primarily clustered with their respective tissues, with spinal cord tissues forming a larger cluster, with the CBL and cortices forming another (**Fig. 4C**).

**Figure 4.**
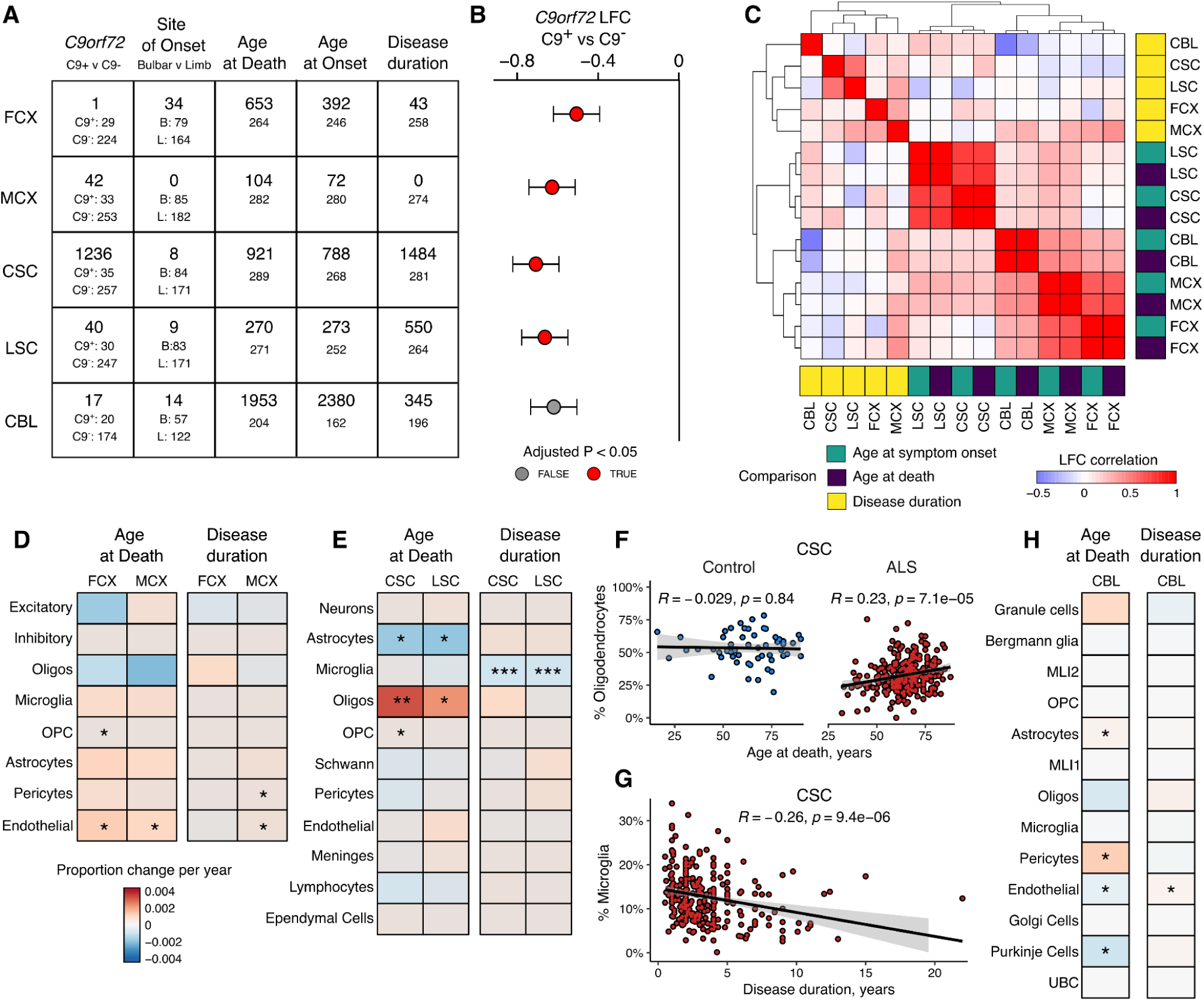
| Gene expression associations with ALS clinical traits. **A**) Summary of all DEGs observed in each comparison within ALS patients in each tissue. Larger numbers refer to DEGs at FDR < 0.05, smaller numbers refer to numbers of samples tested in each comparison. **B**) *C9orf72* is downregulated in C9-ALS in all 5 tissues, plotted as log_2_(fold-change) and standard error. **C**) Pairwise Pearson correlation between DEGs observed in age of onset, age of death, and disease duration between the five tissues. **D-F**) Cellular composition differences associated with age of death and disease duration in cortex (**D**), spinal cord (**E**), and cerebellum (**H**). P-values from linear regression followed by Bonferroni adjustment for the number of cell-types tested. *** P < 0.001; ** P < 0.01; * P < 0.05. **F**) Correlation of estimated proportion of oligodendrocytes in the cervical spinal cord with age at death in controls (blue) and ALS cases (red). **G**) Correlation of estimated microglia proportion in the cervical spinal cord with disease duration. Correlation coefficients and P-values from a two-sided Pearson correlation test. LFC: log_2_(fold-change). OPC: Oligodendrocyte precursor cells; MLI1: Molecular layer interneurons 1; UBC: Unipolar brush cells.

We compared the overlap of differentially expressed genes (DEGs) across clinical categories and performed GO and KEGG enrichment analyses on both unique and shared DEGs across brain and spinal cord regions (**Table S16)**. Due to the limited number of detected DEGs (adjusted p-value < 0.05) and low overlap in some clinical categories, we restricted this analysis to disease duration and age at death. In the age at death category, stratifying significant DEGs by direction of change (upregulated vs. downregulated) revealed that most genes were unique to individual tissues, particularly in the CBL, CSC, and FCX. However, we observed a set of 122 upregulated genes shared between the CSC and LSC enriched in the “Nervous System Development” GO term (**Fig. S8**). In contrast, the disease duration category showed a different pattern: approximately 337 downregulated genes were shared between the CSC and LSC, enriched in “Immune Response” GO term and “Lysosome” KEGG term (**Fig. S8**). Notably, unlike the age at death comparison, FCX showed very few DEGs associated with disease duration (**Fig. 4A**).

We then estimated cell-type proportion changes with age of death and disease duration. In the cortical regions, both age at death and duration identified an increase in the proportion of endothelial cells (**Fig. 4D**). In the spinal cord regions, we observed a significant increase in oligodendrocyte proportions and a decrease in astrocyte proportions with age of death (**Fig. 4E**). We observed that the relationship between age and oligodendrocyte proportions was not observed in the controls (**Fig. 4F; Fig. S9**). Additionally in both spinal cord sections we identified an increase in microglial proportions in shorter disease duration, replicating what we previously observed^6^ (**Fig. 4G**). Although the CBL exhibited weaker overall signals, we detected an increase in pericytes and a decrease in Purkinje cells with increasing age at death (**Fig. 4H**).

### Differential splicing analysis across tissues

Alternative mRNA splicing is known to be perturbed in ALS, both due to changes in cell states and specifically linked to TDP-43 loss-of-function pathology. We performed differential splicing (DS) analyses on the bulk tissue RNA-seq with Leafcutter^51^, which first identifies overlapping clusters of introns that make up a local splicing phenotype and computes differential usage between groups and associations with continuous clinical variables. DS was performed between ALS cases and non-neurological controls across all sampled tissues. At an FDR of 0.05, the number of DS clusters per tissue were as follows: motor cortex (528), cervical spinal cord (1,919), lumbar spinal cord (2,075), frontal cortex (1,407) and cerebellum (103), (**Fig. 5A**), full results are presented in **Tables S17-22**. As with the differential expression analysis, the discovery rate was strongly correlated with the effective sample size (**Fig. S10A**). Pair-wise clustering of cell types by ΔPSI values showed the expected grouping between cortical and spinal cord tissues (**Fig. 5B**).

**Figure 5.**
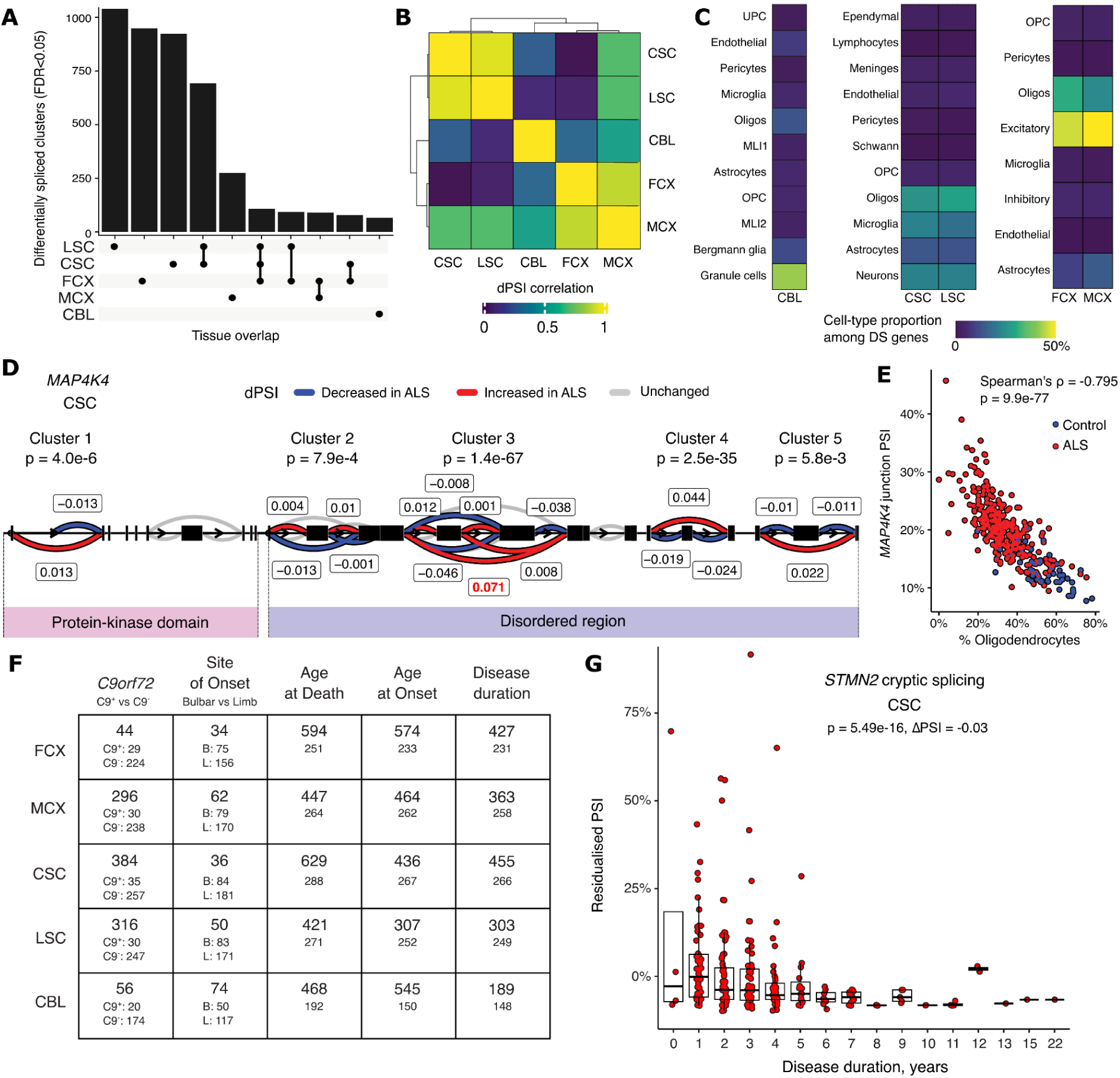
| Differential mRNA splicing analysis reveals differences across tissues. **A)** Overlap of differentially spliced clusters between the five major tissues, limited to the top 9 intersections. **B)** Pearson correlation of representative intron ΔPSI scores for significant DS clusters between the five major tissues with a hierarchically clustered dendrogram. **C)** Proportion of cell-type specific marker genes from tissue-matched references^47^–49 in DS cluster annotations. **D)** Transcript splicing visualization of *MAP4K4* with 5 differentially spliced clusters. The largest effect size intron is labelled in red. **E**) This *MAP4K4* intron negatively correlates with % oligodendrocytes in the cervical spinal cord. **F)** The number of DS clusters for each tissue and comparison within ALS patients. **G)** Cryptic splicing of *STMN2* is associated with disease duration in ALS patients in the cervical spinal cord. Box plots show the median and the first and third quartiles of the distribution, with whiskers extending to 1.5 times the interquartile range.

To link differential splicing to specific cell-types, we used a marker gene approach using the tissue-specific snRNA-seq references to match each gene to its most specific cell-type. Analysis of marker genes for cell types relevant to each tissue revealed clear differences, notably the enrichment of marker genes for granule cells being differentially spliced in the CBL, whereas in cortical regions differential splicing was enriched in markers for neurons and oligodendrocytes, and in the spinal cord splicing was seen in microglia, neurons, oligodendrocytes and astrocytes (**Fig. 5C**).

Our approach identifies both known and novel introns not present in gene annotations. While most introns in differentially spliced clusters were annotated, varying proportions were either fully or partially un-annotated, ranging from 33.47% fully annotated in the CBL to 73.99% in the LSC. Between tissues, genes that were differentially expressed and those that were differentially spliced represented a partially overlapping subset. The lowest overlap was in the CBL (at 20.39%), and the highest in the LSC (63.61%) (**Fig. S10B**). To uncover the potential for differentially spliced junctions to contribute to nonsense-mediated decay (NMD), we utilised a conservative annotation-based approach, as well as a novel annotation-free approach to quantify unproductive splicing^52^. For the annotation-based approach, we assigned NMD potential to junctions that were exclusively part of annotated NMD-inducing transcripts, and noted their direction of effect between cases and controls. We assigned 811 unique NMD-inducing junctions, and observed that only 30.95% of these junctions had a positive ΔPSI (more often used in ALS patients), with the remaining majority being more commonly utilized within the controls. This effect was consistent across all tissues except for the CBL, for which these junctions were evenly utilized between cases and controls. This was also consistent within *de-novo* called NMD junctions (**Fig. S10C**).

For each tissue, a gene-set enrichment analysis was performed using GO biological processes and Reactome pathways on genes for which differential splicing was observed. At FDR<0.05, significant enrichments were observed in the frontal and motor cortex, as well as the cervical, lumbar, and thoracic spinal cord (**Fig. 5C**). The most significantly overrepresented processes between cases and controls were for “actin filament organization” in the LSC (p=1.24e-12) and across age of onset of ALS “modulation of chemical synaptic transmission” in the MCX (p=4.24e-14). Full results are presented in **Tables S29-31**. We were able to observe known examples of transcript splicing events present in ALS, including the TDP-43-associated cryptic exon in *STMN2*^21,53,54^, which we only observed in ALS patients, and predominantly in the LSC (p=2.54e-36) and CSC (p=2.83e-27). We were unable to observe the previously reported cryptic exon in *UNC13A*^22,23^ due to the number of spliced reads falling below our thresholds for cross-tissue analysis. We note that a recent study including this cohort’s bulk RNA-seq data using a targeted approach confirmed the presence of multiple cryptic splicing events across ALS and FTD tissues^55^. We highlight splicing changes in *MAP4K4,* a MAP kinase previously associated with motor neuron degeneration^56^. *MAP4K4* DS events were among the most significant across three tissues affected in ALS (CSC, LSC, and FCX), where 5 distinct splicing regions were differentially used across the gene (**Fig. 5D**). These differentially spliced clusters occurred within coding exons over both protein kinase domains and highly disordered regions. The largest effect size intron in CSC (ΔPSI=7.1%) was in cluster 3 where it is predicted to skip two coding exons in the disordered region. As our cell-type analysis predicted *MAP4K4* to be highest expressed in oligodendrocytes, we correlated the splicing of cluster 3 with the predicted oligodendrocyte proportion in CSC, observing a strong negative correlation (Spearman’s rho = -0.80) (**Fig. 5E**). While none of the *MAP4K4* splice junctions result in an annotated NMD transcript, Cluster 1 was predicted to be unproductive.

Within ALS patients, the continuous clinical variables age at death, age at symptom onset, and disease duration were rescaled and input into a differential splicing model per tissue. At an FDR of 0.05, this resulted in between 307 (LSC) and 574 (FCX) differentially spliced clusters for age of onset, 427 (LSC) and 594 (FCX) for age of death, and between 189 (CBL) and 455 (CSC) for disease duration (**Fig. 5F; Tables S23-28**). Comparing *C9orf72* repeat expansion carrier status, we observed alternative splicing in *C9orf72* for one cluster spanning the first and second exons within the FCX, MCX, and CBL. Large differences between *C9orf72* carriers were also observed in other genes, including the alternative usage of the last exon in the calcium binding-protein *S100B* (p=4.78e-25, max ΔPSI=28.65%), and the ubiquitin-activating enzyme *UBA1* (p=9.76e-28, max ΔPSI=23.3%). Similarly to gene expression, we observed strong overlaps between the genes associated with age of onset and age at death, and a distinct set of genes associated with disease duration (**Fig. S10D**). We observed the same *MAP4K4* splicing cluster 3 associated with disease duration in FCX. We also observed the *STMN2* cryptic exon as negatively associated with disease duration in the CSC and the LSC (**Fig. 5G**). This potentially arises through its proxy status for levels of detectable TDP-43 pathology^21^. Patients with long disease duration may have lower TDP-43 pathology, or due to prolonged time on mechanical ventilation may have lost all TDP-43-containing motor neurons in the spinal cord at the point of death.

### Common variant associations with gene expression and splicing

We then integrated the transcriptomic data generated from each tissue with the paired genotypes generated from WGS to map common variant associations with gene expression (expression quantitative trait loci; eQTLs) and mRNA splicing (splicing quantitative trait loci; sQTLs) using all SNVs and SVs with a minor allele frequency > 0.01, totalling 8,702,972 SNPs and 12,068 SVs. We mapped eQTLs and sQTLs in 11 tissues, splitting the motor cortex (MCX) into three distinct regions: medial motor cortex (MMC), lateral motor cortex (LMC) and unspecified (UMC). In line with previous studies^6,10^, we found that QTL discovery rate, expressed as the percentage of genes tested with a lead association at q value < 0.05, depended highly on sample size, with eQTL discovery being consistently higher than for sQTLs (**Fig. 6A; Table S32**). Due to low sample size impeding QTL discovery, we excluded the hippocampus (HIP), occipital cortex (OCX), temporal cortex (TCX), and thoracic spinal cord (TSC) from downstream analysis. We tested pairwise sharing across tissues using Storey’s π_1_ statistic, and observed high rates of sharing (π_1_ > 0.9) between LSC and CSC, as well as between FCX and MCX samples (**Fig. S11; Table S33**). As previously observed^10^, sharing was higher across all tissue pairs for sQTLs than for eQTLs.

**Figure 6.**
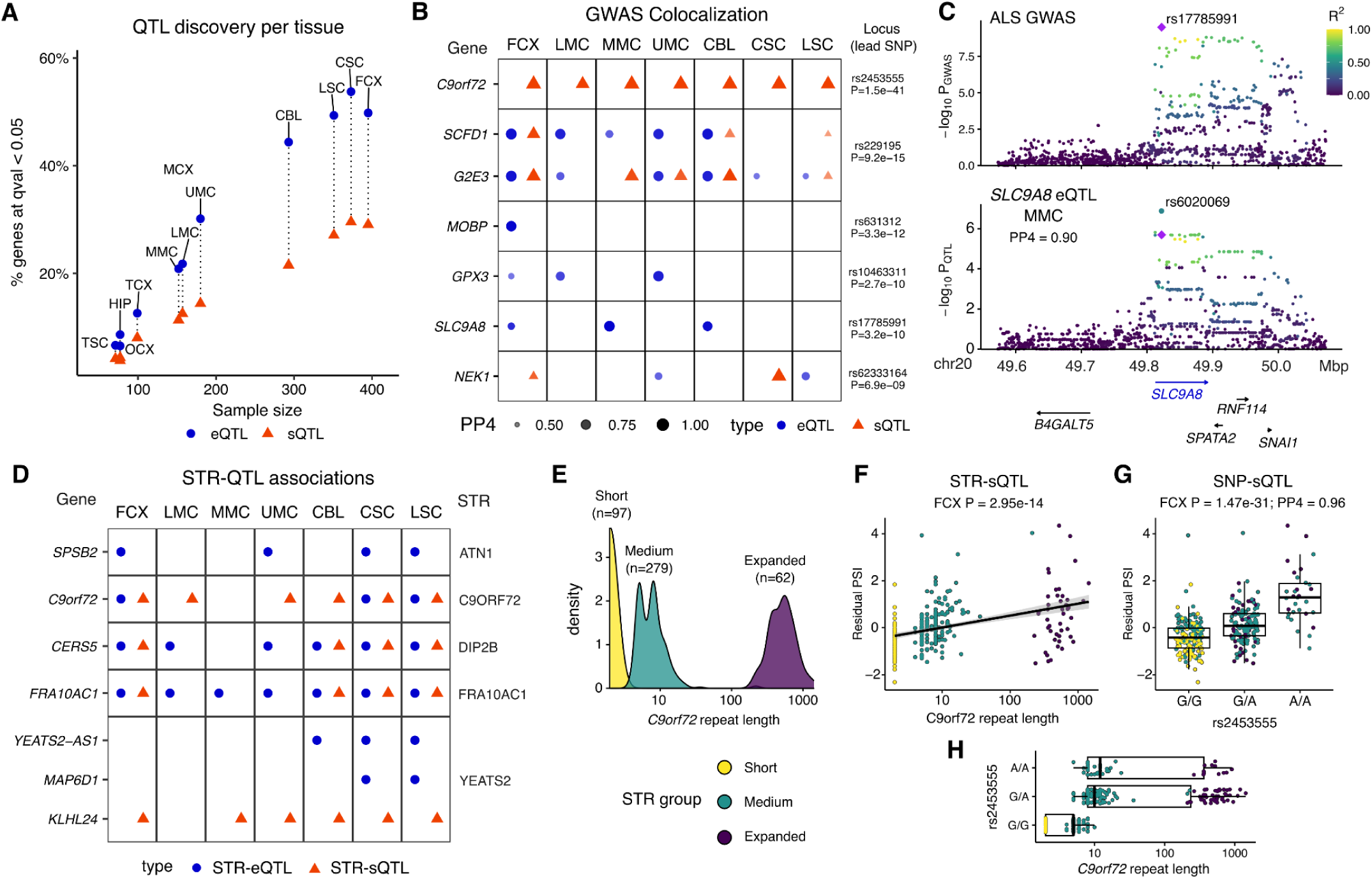
| Common SNPs and STRs are associated with expression and splicing across tissues. **A**) QTL discovery rate for eQTLs (circles) and sQTLs (triangles) in each tissue. **B**) Colocalization of QTLs with ALS GWAS^1^ loci. Only QTL genes with a posterior probability of colocalization (PP4) > 0.9 in at least one tissue are plotted. **C**) The GWAS locus at chr20 (upper panel) colocalizes with an eQTL for *SLC9A8* in the medial motor cortex (MMC; lower panel) at PP4 = 0.90. The lead GWAS SNP rs17785991 (purple diamond) and lead eQTL SNP rs6020069 are close together and overlap the 5’ end of *SLC9A8*. SNPs coloured by LD with rs17785991 in Europeans. **D**) Significant STR-QTL associations with expression (STR-eQTL) and splicing (STR-sQTL). Only STR loci associations found in at least two tissues are plotted. **E**) Distribution of C9orf72 repeat lengths across 438 unique donors. The Gaussian mixture model groups donors into short, medium and expanded repeat lengths for STR-QTL mapping. **F**) The *C9orf72* STR is associated with increased splicing of intron chr9:27567164-27573708 in FCX samples. **G)** The lead GWAS SNP rs2453555 associates with the same intron as an sQTL, which colocalizes with the GWAS locus (PP4=0.96). **H**) The GWAS lead SNP minor allele tags the expanded C9orf72 STR. PSI: percent splicing inclusion. Box plots show the median and the first and third quartiles of the distribution, with whiskers extending to 1.5 times the interquartile range.

We next performed genetic colocalization between our QTL catalogue and the latest ALS GWAS, which identified 15 loci at genome-wide significance (P < 5e-8) in a meta-analysis of European and Asian cohorts^1^. We identified at least one gene colocalizing through expression or splicing at 6 risk loci: *C9orf72, SCFD1/G2E3, MOBP, GPX3, SLC9A8,* and *NEK1* (**Fig. 6B, Table S34**). The top association was seen in *C9orf72,* where a common variant that tags the repeat expansion is associated with sQTLs that alter the balance of splicing between two alternative promoters^6^, which we observe in all 7 tissues. We observed colocalization for expression and splicing in both *SCFD1* and *G2E3* at the same locus, suggesting potential co-regulation of both genes by the same causal SNPs, or two potentially independent effects that cannot be untangled in our data. For the four remaining loci we nominate a single gene, with eQTLs for *MOBP, GPX3,* and *SLC9A8,* and sQTLs for *NEK1.* We highlight the eQTL observed for solute carrier family 9 member A8 *SLC9A8* (**Fig. 6C**). The lead GWAS SNP rs17785991 is 189bp from the lead eQTL SNP for *SLC9A8* in the MMC and the two variants are in moderate (R^2^ = 0.45) linkage disequilibrium (LD) in a European reference panel. The minor T allele of rs17785991 is associated with increased *SLC9A8* expression. Both variants sit within a block of high LD SNPs in the first intron of *SLC9A8*, suggesting a potential cis-regulatory mechanism on *SLC9A8* expression in the motor cortex underlies its association with ALS risk.

### Clinical STR associations with gene expression and splicing

We then used the clinical set of 68 STRs estimated by ExpansionHunter in the PCR-free WGS samples to classify donors into short, medium, and long STR lengths. We associated STR groups with gene expression and splicing of the overlapping genes, adjusting for the same technical and genetic variation as our e/sQTL analysis. At a false discovery rate of 5% we identified 13 STRs associated with gene expression and 10 associated with mRNA splicing, with 6 STRs associated with both, including *C9orf72, DIP2B, and FRA10AC1*. We present the 5 STRs with associations replicated in at least two tissues (**Fig. 6D**; full results in **Fig. S12** and **Table S35**). Notably, no association with gene expression or splicing was found for *ATXN2*, the other clinical repeat expansion associated with ALS^57^. The ataxin family member *ATXN3* was associated with *ATXN3* expression only in the FCX. We were unable to replicate our previously observed association of *ATXN3* STR length with splicing in the spinal cord^6^.

Our STR grouping across the 438 donors with paired RNA-seq identified 62 donors with an expanded *C9orf72* hexanucleotide repeat (**Fig. 6E**). Here we show that the expanded *C9orf72* allele is associated with increased splicing of the *C9orf72* intron chr9:27567164-27573708 (hg38), which splices the exon upstream of the repeat (exon 1a) into exon 2 (**Fig. 6F**). This same intron is driving the colocalized SNP-sQTL, where the minor allele of the GWAS lead SNP rs2453555 is also associated with increased splicing (**Fig. 6G**). Comparing the STR to the lead SNP, we observe that the lead SNP minor allele is likely tagging the expanded STR through linkage disequilibrium (**Fig. 6H**).

### Rare variant outlier analysis

To prioritize rare variants that may act on gene expression, we then identified genes that have an excessive degree of allele-specific expression (ASE), so-called ASE outliers, using the ANEVA-DOT method^58^ (**Fig. 7A**). This approach determines the estimated genetic variation in the dosage of each gene in a population, allowing for the identification of genes where each individual is a population outlier for ASE, suggesting the presence of a heterozygous variant with a particularly strong *cis*-regulatory effect. We performed ASE analysis across 647 individuals (375 ALS cases, 107 controls, 165 other neurological disorders) that had paired WGS and RNA-seq in at least one tissue. For the individuals who had RNA-seq data for multiple tissues, we calculated π_1_ estimates to determine the degree of individual-outlier tissue sharing (**Fig. 7B**)^59^. There was high sharing of the outlier status between the tissues within individuals, particularly between cortical samples (π_1_ = 0.89) and spinal cord samples (π_1_ = 0.85), with the CBL showing modest similarity to the cortical/spinal cord regions (π_1_ = 0.75 and 0.59 respectively). Given the overall similarities of ASE across tissues within individuals, we conducted a meta-analysis across all tissues. Each individual in the meta-analysis had a median of 91 ASE outlier genes (**Fig. S13A**).

**Figure 7.**
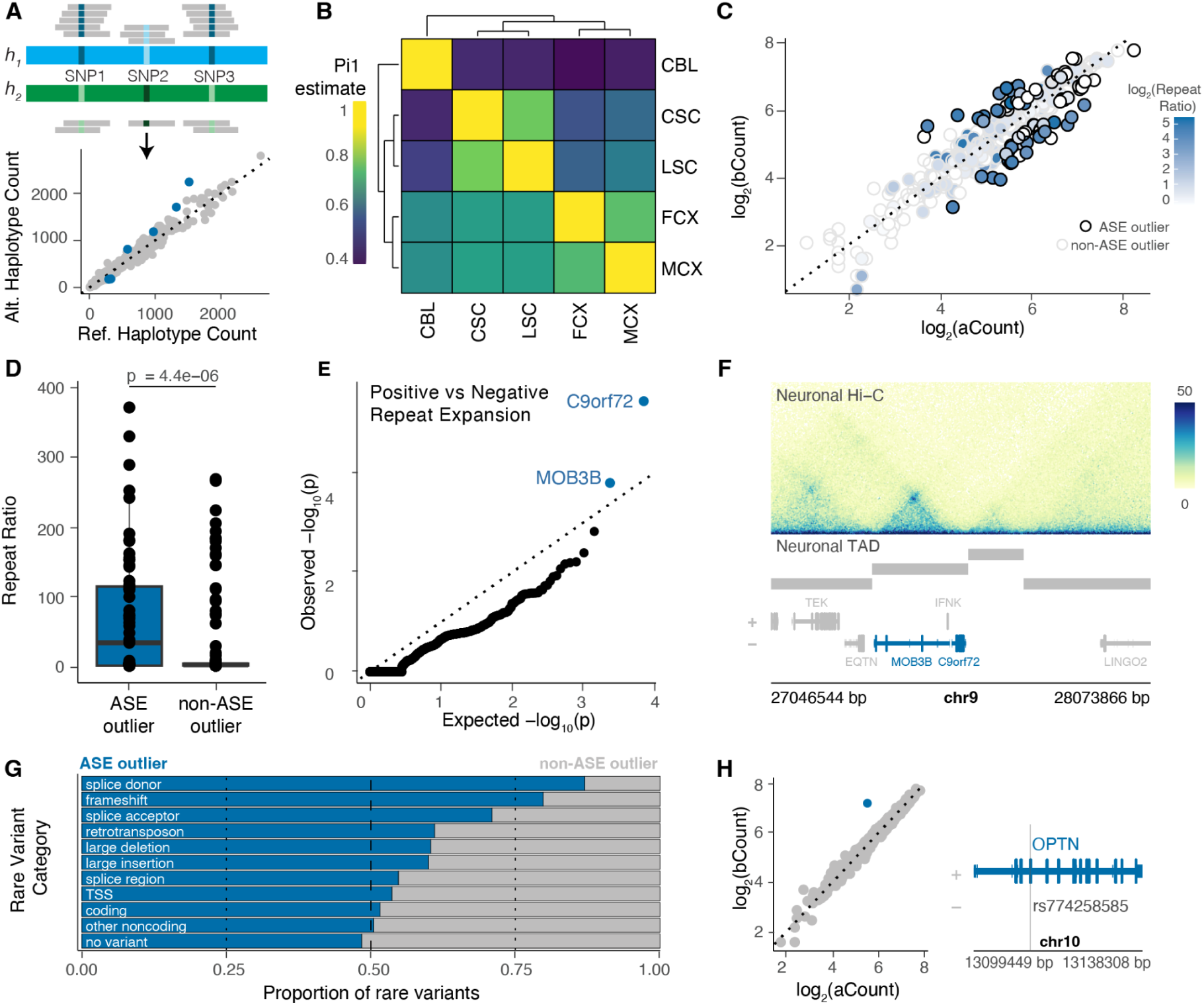
| ASE outliers are enriched for *C9orf72* repeat expansions and rare variants associated with altered gene expression. **A)** Schematic of ASE analysis. RNA-seq and WGS data are integrated to assign reads to different haplotypes for each gene (top). For each gene, the counts assigned to each haplotype (reference and alternative) can be plotted, with ASE outliers shown in blue (bottom). **B)** Heatmap showing the π_1_ estimate of tissue similarity across the five major tissue types with dendrograms showing clustering. Values are not symmetrical across the diagonal as π_1_ = Pr(ASE outlier in tissue *i* given ASE outlier in tissue *j*). **C)** Scatterplot showing the log_2_(haplotypeA) counts on the x-axis and log_2_(haplotypeB) counts on the y-axis of *C9orf72* for all individuals. Black and gray outlines denote individuals that are ASE outliers and non-ASE outliers respectively. The color of the point corresponds to that individual’s *C9orf72* repeat ratio. **D)** Boxplot showing the median and the first and third quartiles for the *C9orf72* repeat ratio for ASE outliers and non-ASE outliers. P-value was calculated from a Wilcoxon Rank Sum Test. **E)** Quantile-quantile plot showing genes enriched for being ASE outliers in individuals positive versus negative for the *C9orf72* repeat expansion. P-values were calculated from one-sided Fisher’s exact test. **F)** Neuronal Hi-C of the *C9orf72* and *MOB3B* region at 5-kilobase resolution. TAD ranges are shown below by gray bars. + and - represent the plus and minus strands respectively. **G)** Barplots showing the proportion of ASE outliers (blue) and non-ASE outliers (gray) with various types of rare variants (MAF < 0.01) Dotted lines represent proportions of 0.25, 0.5, and 0.75. **H)** Scatter plot showing the log_2_(haplotype A) vs log_2_(haplotype B) counts for *OPTN*, with individuals significant for ASE shown in blue (left). Gene tracks showing the location of frameshift mutation rs774258585 (right). + and - represent the plus and minus strands respectively. Box plots show the median and the first and third quartiles of the distribution, with whiskers extending to 1.5 times the interquartile range.

From the meta-analysis, 548 individuals had detectable *C9orf72* allelic expression, 78 of which were ASE outliers for *C9orf72* (**Fig. 7C**). We then analyzed the *C9orf72* repeat counts, and defined the repeat ratio as the ratio between the repeats on each haplotype. Individuals that were ASE outliers for *C9orf72* had a higher median repeat ratio (34 vs. 2.5, p=4.449x10^-6^) than individuals that were non-ASE outliers (**Fig. 7D**). This indicates that the repeat expansion affects ASE of *C9orf72*. There were several individuals that are not ASE outliers but are positive for the *C9orf72* repeat expansion (n = 18), but these individuals had fewer ASE outliers (p = 0.02, **Fig. S13C**) and lower coverage at heterozygous haplotypes (p = 0.00036, **Fig. S13D**) per individual than those individuals negative for the repeat expansion. This suggests that these samples are under-powered to identify ASE outliers and are likely false negatives. We also observed individuals that are ASE outliers but negative for the *C9orf72* repeat expansion (n = 13) (**Fig. S13B**). This set of *C9orf72* ASE outliers that are negative for the repeat expansion had the weakest p-values (12 out of 13 individuals had an FDR greater than the median of 2.97 x 10^-9^, **Fig. S13G**), and had the weakest absolute fold-change between their haplotypic counts per allele (aFC, all 13 samples were less than the median of 0.83, **Fig. S13E-H**). This all supports that these individuals are the weakest *C9orf72* ASE outliers.

While *C9orf72* was the most enriched gene as an ASE outlier in the repeat expansion group, the *MOB3B* gene was also enriched (**Fig. 7E**). Interestingly, *MOB3B* is the gene immediately downstream of *C9orf72* on the same strand and is an ASE outlier specifically in the CBL (**Fig. S13I**). Since the CBL is composed mostly of neurons^60^, we examined this region in neuronal Hi-C data and found that these two genes reside within the same topologically associating domain (TAD) (**Fig. 7F**)^61^. Together these results highlight an interesting case of disease-associated ASE where genetic variation disrupts both the gene where the STR resides and the nearest downstream gene.

In ALS case vs control analysis of ASE outlier status, *C9orf72* was also enriched, along with genes that have been previously implicated in other neurodegenerative disorders, such as *COX6A1* and *TMPRSS5* (**Fig. S13J**)^62,63^. It has been previously reported that ASE outliers are enriched for rare variants (RVs) that often trigger nonsense-mediated decay, such as frameshift mutations and premature stop codons^58,64,65^. We identified RVs (MAF < 0.01) that are nearby both ASE outliers and non-ASE outliers across the cohort and determined the proportion of RVs present for each category in ASE outliers and non-ASE outliers. For many categories of RV annotations, we found that they had higher proportions in ASE outliers than non-ASE outliers (**Fig. 7G**). In particular, frameshift and splice donor variants were the most prominent, with a proportion of > 0.75 in ASE outliers (**Fig. 7G**). An interesting example of this exists for *OPTN*, a gene that has been previously implicated in ALS. One ALS donor in our cohort has a rare frameshift mutation (rs774258585) that increases the risk of ALS (**Fig. 7H**). This individual is the only ASE outlier for *OPTN* in the entire cohort (**Fig. 7H**). In another example, four individuals (1 ALS/FTD, 3 FTD) were ASE outliers for the *GRN* gene in the frontal cortex, and all of these individuals had the frameshift mutation rs63751057 (**Fig. S13K**). Together, these two examples suggest a potential mechanism whereby the frameshift mutations drive ASE in their respective genes, increasing the risk of ALS.

## Discussion

We present the definitive and complete release of the NYGC ALS Consortium dataset, the largest multi-tissue genomic resource for ALS assembled to date, and demonstrate its capacity to yield biological and genetic insights. The immediate scientific value of open data sharing is demonstrated by the multiple independent groups who have already generated findings from this resource prior to this publication, including the identification of new ALS risk genes^2,3^, TDP-43 cryptic splicing biomarkers^21–23,25–27^ and ALS molecular subtypes^7–9,20^. We present our analyses as a foundation and invitation for the community to build upon in their own studies.

We note that the consortium’s open inclusion model prioritised scale and speed of data generation. As a consequence, the cohort has limited ancestral genetic diversity and a relative paucity of non-neurological controls, constraining case-control genetic association analyses within the cohort alone. Future efforts should complement this resource with targeted recruitment to improve ancestral representation. Additionally, the cohort focused on donors with a primary diagnosis of ALS, as we analyzed the RNA-seq from the FTD donors in the cohort in an earlier study^28^. A future joint analysis spanning the full ALS-FTD spectrum, paired with direct measurement of TDP-43 pathology burden, would provide a more complete picture of shared and distinct molecular pathology across TDP-43 proteinopathies.

Our WGS variant catalogue identified likely pathogenic mutations in 15.6% of ALS patients across known ALS genes, capturing the full spectrum of small variants and short tandem repeats. This figure will increase as genetic association studies continue to expand the catalogue of confirmed ALS mutations^2^ and as long-read DNA sequencing enables more accurate genotyping of complex SVs and STRs^66^. A practical limitation of our cohort is the underrepresentation of rare mutation carriers in the matched RNA-seq data: *TARDBP*, *FUS*, and *NEK1* carriers in particular have few or no transcriptomic samples. Future cohort construction should prioritise matched transcriptomic sampling of these carriers.

Transcriptomic analysis across five major tissues revealed that ALS, despite its heterogeneous clinical presentation, converges on a shared end-stage molecular pathology. The high concordance between frontal and motor cortex, and between cervical and lumbar spinal cord, suggests that by the point of tissue collection, regional differences in symptom onset have been subsumed by common disease processes, a conclusion reinforced by the lack of differential gene expression or splicing between donors with limb or bulbar onset. The most therapeutically actionable finding from our splicing analysis is the identification of *MAP4K4* as a differentially spliced gene across ALS tissues and its splicing association with oligodendrocyte proportions in the spinal cord and with disease duration in the frontal cortex. *MAP4K4* encodes a kinase with established roles in neuroinflammation and neurodegeneration, and existing small-molecule inhibitors have shown promise in preclinical models of motor neuron disease^56^. Our results enable the field to characterise *MAP4K4* splicing at a cell-type level using targeted approaches in tissues and biofluids.

The observation of increased lymphocyte proportions in ALS spinal cord, and the correlation of *STMN2* cryptic splicing with disease duration in spinal cord tissue, are findings that similarly require validation and mechanistic follow-up in independent cohorts and experimental systems. The cerebellum warrants particular attention as a regional outlier across all our transcriptomic analyses, showing the fewest differentially expressed and spliced genes of any tissue examined. This is paradoxical given reports of dipeptide repeat pathology in cerebellar tissue in C9orf72-ALS^67–69^. Whether the cerebellum is genuinely spared from the transcriptomic consequences of ALS pathology, or whether its distinct cellular composition renders bulk RNA-seq an insensitive readout, is an open question our resource is well-positioned to help resolve.

Integrating the genetic and transcriptomic data, we find that *C9orf72* repeat expansions consistently associate with reduced total *C9orf72* expression and altered splicing of the exon 1a isoform, confirmed by three orthogonal approaches: common variant sQTL mapping, direct modelling of the repeat length, and allele-specific expression. The enrichment of *C9orf72* expansion carriers in the cohort (85/695; 12.2% of all donors with matched RNA-seq and WGS) drove this result’s statistical power. Beyond the *C9orf72* gene, genome-wide transcriptomic comparison between *C9orf72* carriers with non-carriers identified few differentially expressed genes or splicing events, consistent with end-stage pathology obscuring mutation-specific effects. The resource enables future studies to address this with larger genetic subgroup sample sizes as cohorts grow and through integration with single-cell data that can isolate mutation-specific effects in individual cell types.

Our dataset complements emerging single-nucleus and spatial transcriptomic approaches. These methods offer cellular resolution that bulk RNA-seq cannot provide, but currently at sample sizes an order of magnitude smaller than achieved here. Bulk tissue RNA-seq captures population-level transcriptomic signals but cannot distinguish true cell-type composition changes from cell-state shifts, and is limited in its ability to detect rarer populations including upper and lower motor neurons. Differential splicing analysis from single-cell data remains technically challenging^70^, and our study establishes the groundwork for future single-cell and spatial transcriptomic studies of the same tissues to resolve these questions with cellular precision. Our resource’s large patient numbers and tissue breadth enable clinical subgroup analyses that small single-cell cohorts cannot support, whereas single-cell data from the same tissues can resolve the cellular attribution of the population-level signals. We propose that integration of our immune infiltration and glial activation findings with targeted single-cell profiling of spinal cord immune populations will clarify their therapeutic relevance.

In summary, the NYGC ALS Consortium dataset provides a foundational resource for the field, enabling integration of genetic variation with the molecular and clinical features of ALS at a scale and tissue breadth not previously achievable. We anticipate that its greatest contributions will come from studies not yet conceived.

## Methods

The NYGC ALS Consortium samples presented in this work were acquired through various IRB protocols from member sites and the Target ALS postmortem tissue core and transferred to the NYGC in accordance with all applicable foreign, domestic, federal, state, and local laws and regulations for processing, sequencing, and analysis. The Biomedical Research Alliance of New York (BRANY) Institutional Review Board serves as the central ethics oversight body for NYGC ALS Consortium. Ethical approval was given and is effective through 06/18/2026. All necessary patient/participant consent has been obtained and the appropriate institutional forms have been archived.

### Whole Genome Sequencing

DNA was extracted from whole blood or flash frozen post-mortem tissue (**Table S1**) using QIAamp DNA Mini Kit (QIAamp #51104 and QIAamp#51306, respectively) according to the manufacturer’s recommendations. Incoming nucleic acid samples are quantified using fluorescent-based assays (PicoGreen or Qubit) to accurately determine whether sufficient material is available for library preparation and sequencing. DNA sample size distributions were profiled by a Fragment Analyzer (Advanced Analytics) or BioAnalyzer (Agilent Technologies), to assess sample quality and integrity. Samples with a Genomic Quality Number (GQN), according to the Fragment Analyzer results, below 2.5 were dropped from the study.

We sequenced 4,828 DNA samples, including 2,001 females and 2,827 males. Over the duration of the study, samples were sequenced on HiSeq X or NovaSeq 6000 machines using Illumina TruSeq PCR-free, Illumina TruSeq Nano PCR+ or Roche KAPA Hyper PCR+ library preparation kits (see **Methods Table 1**). 82 WGS samples failed sample QC (% chimeric reads > 5% and AT dropout > 8.75) and were excluded from downstream analysis, resulting in 4,746 WGS samples (1,974 females and 2,772 males) that were included in small variant calling and the integrated SNV/INDEL/SV callset.

Whole genome sequencing (WGS) libraries were prepared using the Illumina TruSeq DNA PCR-free Library Preparation Kit in accordance with the manufacturer’s instructions. Briefly, 1ug of DNA is sheared using a Covaris LE220 sonicator (adaptive focused acoustics). DNA fragments undergo end-repair, bead-based size selection, adenylation, and Illumina sequencing adapter ligation. If at least 800ng were present, the PCR-free protocol was used. For samples with under 800ng, the Illumina TruSeq DNA Nano Library Preparation Kit in accordance with manufacturer’s instructions. Briefly, 100ng of DNA is sheared using the Covaris LE220 sonicator (adaptive focused acoustics). DNA fragments undergo end-repair, bead-based size selection, adenylation, and Illumina sequencing adapter ligation. Ligated DNA libraries are enriched with PCR amplification (using 8 cycles). For samples prepped with Kapa Hyper, 100ng of DNA was used as input, with PCR amplification (using 4 cycles). Picogreen is used to measure the total amount of DNA in the prepared library. Quantitative PCR (qPCR) uses specific oligos complementary to Illumina’s TruSeq adapters to measure the amount of adapter-ligated DNA (ligation efficiency) that is compatible with sequencing. For WGS samples, an aliquot of DNA is separately aliquoted from the submission tube upon receipt for SNP array genotyping to determine DNA integrity and identity ahead of sequencing. The genotyping results are also checked for gross sample contamination and can reveal other forms of poor sample quality prior to sequencing. This SNP array was also used to confirm that duplicate patients were not being introduced into the study.

### Small variant calling

WGS data for 4,746 samples was processed on an NYGC automated pipeline^38^ that follows the recommendations outlined in the functional equivalence standard developed for the Centers for Common Disease Genomics project^71^. Paired-end 150bp reads were aligned to the GRCh38 human reference using the Burrows-Wheeler Aligner (BWA-MEM v0.7.15), marking of duplicate reads using Picard tools (v2.4.1; see **URLs**), and base quality score recalibration (BQSR) via Genome Analysis Toolkit (GATK)^29,72^ v3.5. In addition, an in-house short alignment marking (SAM) tool was applied to a subset of 2,304 samples after BWA-MEM. SAM unmaps reads with fewer than 30 bps aligned to the reference–typically arising from cross-species contamination–and is part of the functional equivalence standard^71^. SNV and INDEL calling was performed using GATK HaplotypeCaller v3.5^73^ in GVCF mode, with sex-dependent ploidy settings applied to chromosomes X and Y. Specifically, variant calling on chromosome X was performed with `ploidy=2 in females, ploidy=2 in pseudoautosomal (PAR) regions in males, and ploidy=1 in non-PAR regions in males. Variant calling on chromosome Y was performed only in male samples with ploidy`=1 setting. Genetically inferred sex was computed using an in-house tool, Binest. Binest estimates sex by calculating data density from the BAM index (.bai) in 16,384 bp windows in the reference on sex chromosomes. It then normalizes these values against autosomal medians to provide an estimated copy number for sex chromosomes. We used a cutoff for median data density in bytes for every 16 kbp window on chrY of 1e-5 to assign genetically inferred males and females. Resulting GVCFs were combined in batches of 200 using GATK CombineGVCFs. After merging, discovered variants were jointly genotyped across all 4,746 WGS samples using GATK GenotypeGVCFs producing cohort-level SNV/INDEL VCF. We then assigned a quality tranche to each variant using GATK VariantRecalibrator which uses variant annotations to train the Variant Quality Score Recalibration (VQSR) model using `maxGaussians 8` and `maxGaussians 4` parameters for SNVs and INDELs, respectively. We applied the VQSR model to the joint call set using ApplyRecalibration with tranche cutoffs of 99.8% for SNVs and 99.0% for INDELs.

### Ancestry analysis

Our ancestry pipeline estimates individual genome-wide average ancestries from a set of SNP genotypes using ADMIXTURE^74^, a maximum likelihood-based method. The pipeline takes a gVCF generated by Haplotype Caller as input, runs through a series of processing steps in plink and passes the processed plink output to ADMIXTURE which performs ancestry estimation. The 1000 Genomes phase 3 project samples^75^ were used as the reference populations. The pipeline estimates ancestries for individual samples at the 1000 Genomes defined “super population” level which are: AFR: African, AMR: Admixed Americas, EAS: East Asian, EUR: European, and SAS: South Asian. Samples from the MXL (Mexican Ancestry from Los Angeles USA) and ASW (Americans of African Ancestry in SW USA) populations were excluded from the reference because they might be putatively admixed. The values range from 0-1 to represent the estimated fraction of each population to which the sample belongs.

### Short tandem repeat calling

We called STR expansions in whole-genome sequencing data using ExpansionHunter (EH)^40,76^ v5.0. EH estimates the number of copies of repeated short unit sequences by performing a targeted search through a BAM/CRAM file for reads that span, flank, or are fully contained in each repeat. By combining evidence from multiple read signals, this approach is capable of genotyping repeats at a locus of interest even when the expanded repeat is substantially larger than the read length. The method is able to discover and accurately estimate the size of *C9orf72* repeat expansions containing as many as ∼1000 (∼6Kbp) copies of the motif. Our catalog genotyped repeat expansions across the 4,746 cohort samples in the following 68 clinical STR loci using EH: *ABCD3, AFF2, AFF3, AR, ARX_EIEE, ARX_PRT, ATN1, ATXN1, ATXN10, ATXN2, ATXN3, ATXN7, ATXN8OS, BEAN1, C9orf72, CACNA1A, CBL, CNBP, COMP, CSTB, DAB1, DIP2B, DMD, DMPK, EIF4A3, FGF14, FMR1, FOXL2, FRA10AC1, FXN, GIPC1, GLS, HOXA13_1, HOXA13_2, HOXA13_3, HOXD13, HTT, JPH3, LRP12, MARCHF6, NIPA1, NOP56, NOTCH2NL, NUTM2B_AS1, PABPN1, PHOX2B, PPP2R2B, PRDM12, PRNP, RAPGEF2, RFC1, RILPL1, RUNX2, SAMD12, SOX3, STARD7, THAP11, TBP, TBX1, TCF4, TNRC6A, VWA1, XYLT1, YEATS2, ZFHX3, ZIC2, ZIC3,* and *ZNF713.* This variant catalog was curated from multiple sources including the gnomAD Tandem Repeat Browser^77^ v3.1.2, STRchive^78^, and STRipy^79^; see **URLs**. We excluded the *HOXA13* locus due to challenges in repeat characterization likely arising from the proximity of imperfect GCN motifs and thus resulting in an elevated false-positive rate^79,80^. Trtools (v5.0.2)^81^ mergeSTR was used to merge single sample VCFs into a multi-sample VCF file, and bcftools v1.18 was used to compress and index the newly merged VCF. Genotype distributions of both alleles were plotted (**Fig. S3A**).

Repeat-primed PCR (RP-PCR) assays were used to detect and size repeat expansions for *ATXN2* and *C9orf72* and to validate our EH length estimates. The number of CAG repeats in *ATXN2* was determined in 103 cohort samples using fragment analysis/capillary electrophoresis of fluorescently-labeled PCR products using PCR conditions as previously described^82^. We examined the correlation between *ATXN2* EH repeat length and length estimates from the RP-PCR assay (Figure S3B). Of the 103 samples, 3 have a minor length discrepancy between EH and the RP-PCR estimates: 2 samples differ by 1 repeat length and 1 sample differs by 3 repeat lengths. *C9orf7*2 was genotyped in 240 cohort samples using RP-PCR as previously described^83^. Primers were tagged with a 5’ FAM fluorophore so that fragment size could be measured on capillary electrophoresis. We compared the RP-PCR-determined *C9orf72* expansion status ≥30 repeats (yes/no) to EH.

### Relationship Estimation

We performed IBD analysis on the 4,746 samples using Kinship-based INference for Gwas (KING)^84^ 2.2.3. To perform this analysis, the joint genotyped WGS VCF files were converted to Plink binary files, and only high-quality sites were selected for analysis (genotype call rate >95%, MAF >1%). Analysis was performed on LD pruned sites (r^2^< 0.5). **Fig. S4** highlights the relationships identified in the KING analysis. There are 355 inferred relationships up to the second degree: 91 Parent-offspring (PO), 171 Full siblings, and 93 2nd-degree relatives (2-Deg). The remaining more distant relationships include 124 3rd-degree (3-Deg) relative pairs, 240 4th-degree (4-Deg) relatives and 11,259,166 unrelated (UN) pairs.

### Structural Variant Calling

#### Manta

For every cohort sample, Manta v1.5^32^ was run with default parameters. Both insertions and deletions with a PASS filter flag were pushed to their left-most possible position in the reference and genotyped on the same sample using Paragraph v2.4a^34^ in a ploidy-aware manner with the option -M corresponding to 5 times the average sample coverage. Only variants with a PASS non-reference genotype (0/1, 1/1 or 1) were selected to ensure that variants that didn’t produce non-reference genotypes in the sample where they were discovered were not carried into merging. Prior to that operation, insertions that originally had an incomplete sequence representation, consisting of two disjointed breakends, were transformed by connecting their sequences – extracted from the corresponding INFO field tags LEFT_SVINSSEQ and RIGHT_SVINSSEQ – via a 200 N-character gap to allow genotyping by Paragraph. Insertions and deletions, processed independently, were then clustered based on breakpoint position by connecting all variants where both breakpoints (left and right) are located within 500bp of the corresponding breakpoint of any other variant in the cluster. Then, for each cluster, local haplotypes were reconstructed by padding variants with reference sequence from, and up to, the same reference positions, ensuring a minimum padding of 500bp upstream and downstream of the left-most and right-most breakpoints in the cluster, respectively. These local haplotypes were then aligned using MAFFT^85^ v7.525. The resulting multiple sequence alignments allowed for the separation of variants that didn’t overlap in alignment space into independent clusters and, for each of the resulting subclusters, the selection of the most consensual sequence representation. The resulting set of variants, one per subcluster, were then genotyped on all samples of the cohort using Paragraph to produce two unified call sets – one consisting of insertions and another of deletions, both genotyped across the entire cohort. Non-PASS genotypes as determined by Paragraph, were converted to missing (./. or ., depending on ploidy).

#### Absinthe

For every cohort sample, we ran Absinthe (see **URLs**), which is an insertion caller based on global *de novo* assembly of unmapped and partially mapped reads. Insertions are called by aligning the resulting contigs to the reference and subsequently genotyped using Paragraph v2.4a^34^. Then, using the same process described above for merging Manta SV calls, PASS-filter insertions from all samples were clustered and the most consensual haplotype sequence of each cluster was selected as the representative variant sequence for each locus. The resulting set of variants were then genotyped across the entire cohort using Paragraph v2.4a to produce a unified insertion call set, where non-PASS genotypes were converted to missing.

#### HGSVC-paragraph

Insertions and deletions from v2.0 call set produced by HGSVC^36^ were genotyped with Paragraph v2.4a^34^ across the entire cohort. The resulting single-sample VCFs were combined using BCFtools^86^ to produce a multisample VCF. Like for Manta and Absinthe VCFs, non-PASS genotypes were converted to missing.

#### HGSVC-pangenie

Insertions and deletions from the Pangenie PAV-panel, released as part of the HGSVC v2.0 call set, were genotyped with Pangenie^37^ v2.1.0 across the entire cohort. The individual sample results are filtered to contain only SVs – the PAV-panel consists of SNVs, INDELs and SVs – with a genotype quality of 200 or greater. Filtered per sample calls were merged into a multisample VCF with the same approach as the HGSVC-paragraph results. As in previous analyses, non-PASS genotypes were converted to missing.

#### MELT

We ran MELT^35^ v2.2.2, which has five components: preprocessing, individual analysis, group analysis, genotyping, and VCF creation. Preprocessing is done first for every cohort sample. Next, the individual analysis is performed per Mobile Element Insertion (MEI) type – ALU, LINE1, SVA – on each cohort sample. Following this, per sample results for each MEI type are combined in the group analysis. The combined calls for each MEI type are then genotyped in every cohort sample. Finally, the individually genotyped results are combined into final VCFs per MEI type. Only MEIs with a PASS filter flag were selected. For those, genomic positions reported by MELT were adjusted, so they are consistent with calls produced by Manta, Absinthe and the HGSVC. This was done by shifting them upstream by a number of bases corresponding to the target site duplication sequence length reported for each MEI. Additionally, given that MELT only produces diploid genotypes, these were converted to haploid over non-PAR regions of chromosomes X and Y in males.

#### Ensemble SV call set

SVs from the previously described single-tool call sets were merged into a single call set. This was done independently for insertions and deletions. First, variants from each call set that failed Hardy-Weinberg Equilibrium Chi-square test (p-value <= 10^-10^) and presented more than 10% genotype missingness were filtered out. The remaining variants were then clustered based on genomic position using a strategy similar to that above described for Manta and Absinthe with the difference that, in this case, breakpoints from all variants (left and right separately) in a given cluster must be within the maximum distance (50bp) of those from all other variants in the same cluster and not just at least one. Then, within each cluster, a representative variant was selected based on the following criteria: 1) having a sequence representation; 2) lowest missingness; 3) greater SV length; and 4) discovered on the cohort, opposed to coming from an external call set (HGSVC). The remaining cluster variants are still listed in the INFO field of the representative variant VCF record, along with the corresponding genotype concordance, and non-reference genotype concordance to it. Deletions longer than 1kb were further screened based on the ploidies observed for the different genotypes across the cohort. For a given deletion, ploidy for each sample was calculated by comparing depth of coverage across the deletion’s reference span, computed using Mosdepth^87^ v0.3.3, to the sample’s average. Deletions for which no significant ploidy difference between different genotypes was observed (mean delta < 0.5 or one-sided Wilcoxon Rank Sum test p-value > 0.05) were discarded. Finally, genotypes from 111 samples (29 unconsented and 82 that didn’t pass QC on percentage of chimeric reads and AT dropout), as well as variants that were only present in those samples, were removed. Variants with allele frequency of 0 or 1 were also removed. Variant callset integration and variant filtration

We integrated the 4,746 sample small variant (SNV & INDEL) and structural variant (large DELs and INSs) callsets using bcftools v1.19^86^ `concat` with `--allow-overlaps` option, followed by `bcftools sort`. Upon integration, we annotated all variants with Hardy-Weinberg Equilibrium (HWE) exact test p-value^88^ using the `bcftools fill-tags` plugin. We used `bcftools annotate` to annotate sites with >=80% overlap with low complexity regions (LCR) of the genome (labeled “LCR80” in the INFO field of the VCF), as defined by the low complexity genomic stratification bed file v3.3 from The Genome in a Bottle Consortium^89^ (see **URLs**). Additionally, we defined the following site-level QC metrics in the INFO field of the integrated VCF:

- CR95_QC=PASS/FAIL: PASS, if genotype call rate > 95%.
- HWE_QC=PASS/FAIL: PASS, if HWE exact test p-value > 1e-10.
- LCR80_QC=PASS/FAIL: PASS, if LCR80 annotation is absent, i.e. if a site is outside of low complexity regions or if less than 80% of it overlaps low complexity regions (in case of INDELs).
- Stringent_QC=PASS/FAIL: PASS, if:

- FILTER=PASS (FILTER flag PASS indicates passing VQSR in case of SNV/INDELs; whereas in the case of SVs it indicates meeting the following QC criteria: AC>=1, PFR >=90%, HWE chi-square test p-value >1e-10).
- CR95_QC=PASS.
- HWE_QC=PASS.
- LCR80_QC=PASS. Mendelian error rate by variant type

As described in the Relationship Estimation section, there are 11 genetically-inferred father-mother-child trios among the 4,746 WGS samples. We utilized the presence of trios in our cohort to assess the quality of genotypes called in the integrated callset by computing mendelian error rate (MER) in child samples. MER was defined as a fraction of child sample genotypes that are inconsistent with inheritance patterns out of all non-homREF genotypes in a given child sample. Only sites with non-missing genotypes in child, father, and mother samples were included in MER computation. MER computation was stratified by variant type (SNV, INDEL, SV), minor allele frequency in the full cohort (rare: MAF < 1%, common: MAF ≥ 1%), and variant filtering conditions (i) FILTER=PASS, ii) FILTER=PASS+CR95_QC=PASS+HWE_QC=PASS, and iii) FILTER=PASS+CR95_QC=PASS+HWE_QC=PASS+LCR80_QC=PASS).

### Functional annotations

We used Variant Effect Predictor (VEP) v114.0^39^ with Singularity v4.2.2^90^ to annotate the integrated SNV/INDEL/SV variant calls with functional information. The following options were used when running VEP:

--cache --offline --exclude_predicted --fasta ${fastaReference} --gencode_basic --sift p --polyphen p --hgvs --symbol --numbers --domains --regulatory --tsl --gene_phenotype --pubmed --nearest symbol --pick_allele_gene --af_gnomade --af_gnomadg –vcf --compress_output bgzip --force_overwrite --fork 4 --input_file ${vcf} --output_file ${outdir}/${vcf_prefix}.vep_v114.vcf.gz --custom

`file=${clinvarVCF},format=vcf,short_name=clinvar_20250504,type=exact,overlap_cutoff=0,fie lds=DBVARID%ALLELEID%CLNDN%CLNDISDB%MC%CLNSIG%CLNSIGCONF%CLNREVSTAT%ORIGIN.`

### Annotation of ALS mutations

We appreciate that not all research groups have the technical capabilities of reprocessing the WGS data to identify participants with variants they may want to study. We aimed to create a list of participants who carry variants that are highly likely to be influencing ALS/FTD risk. Using the small variant and structural variant VCFs, we filtered for QC-passing variants with any VEP coding annotation for a list of 154 genes associated with ALS genes and mimics of ALS (**Supplementary Tables**). Variants with ClinVar assertions of benign or likely benign were excluded. Variants were then re-annotated with gnomAD^77^ v4.1 allele frequencies, and those with a global MAF >0.001 were excluded unless they carried a ClinVar assertion of pathogenic or likely pathogenic. Any remaining variants were manually reviewed by the Columbia ALS VUS S.O.S. program methodologies^91^ to modify any ACMG classifications based on case counts from in-house/publicly available ALS databases, comprehensive literature review, and functional effect prediction algorithms. These classifications are intended to identify variants that may be related to the participants disease but should not be confused with standard ACMG classifications. Undoubtedly, there are variants not included in these lists that may have some evidence for causation. Investigators interested in other variants can reprocess and classify the variants themselves. Additionally we screened the EH STR VCF for expansions in the standard panel of expansions for those in the intermediate or pathogenic ranges. STR expansions in the range associated with “reduced penetrance” were classified as likely pathogenic, while those associated with “high or full penetrance” were classified as pathogenic.

Participants were categorized as “ALS single gene” if they: 1) carry a single heterozygous pathogenic or likely pathogenic variant in a known dominant or semi-dominant ALS gene; 2) are homozygous for a pathogenic or likely pathogenic variant in a gene that causes ALS with recessive inheritance; 3) are potentially compound heterozygous for a gene that causes ALS with recessive inheritance. We required that at least one of the two variants had a pathogenic or likely pathogenic classification. We excluded any pair of variants if the haplotype predictor in gnomAD v4.1 showed a high likelihood of being on the same haplotype. Participants were categorized as “Risk factor/Carrier/Intermediate” if they carry a single variant in a gene that is biallelic for ALS or an intermediate EH repeat length in *ATXN2* (31-33 copies) or *C9ORF72* (20-29 copies). Participants were categorized as “Oligogenic” if they carried relevant variants in at least 2 genes related to ALS. Although a handful of participants carry two potentially causative variants, the majority of participants in this group are those carrying a clearly causative variant with a second variant that is best understood as being a carrier in a recessive gene, or a risk factor allele (e.g. ATXN2 intermediate repeat).

Samples annotated as having a pathogenic C9orf72 expansion were either those with PCR-free WGS and an EH call ≥ 30 repeats or PCR+ WGS with RP-PCR validation. This criteria overrode whether the submitting site had called them as having a *C9orf72* expansion due to concerns about false negatives from historical samples. We note that a small number of missense variants in *SETX* are associated with dominantly inherited ALS via a gain-of-function mechanism^92^. These were considered as potentially causative of ALS. Bi-allelic loss-of-function variants in *SETX* are causative of ataxia with ocular apraxia (AOA). Single loss-of-function variants in *SETX* have not been associated with ALS, so these were considered “carriers”. Similarly, although missense variants in *SQSTM1* have been reported in ALS and FTD, they are most strongly associated with Paget’s disease of bone^43^. Thus, potentially pathogenic or likely pathogenic missense variant carriers were considered “mimics” rather than “ALS Single gene.” Bi-allelic loss-of-function variants in *SQSTM1* cause neurodegeneration with ataxia, dystonia, and gaze palsy, childhood-onset^93^, but single loss-of-function variants are not associated with ALS and carriers of these are considered “carriers.”

### RNA sequencing

Total RNA was extracted from flash frozen post-mortem tissue. Trizol/Chloroform extraction method is used, followed by Qiagen RNeasy Mini kit column purification. The column purification step is used to ensure the quality of extracted RNA. Total RNA was quantified using fluorescent-based assays (RiboGreen or Qubit) to accurately determine whether sufficient material is available for library preparation and sequencing. A Nanodrop 2000 reading provides a 260/280 ratio, a generally accepted measure of purity of extracted RNA. RNA sample size distributions were profiled by a Fragment Analyzer (Advanced Analytics) or BioAnalyzer (Agilent Technologies), to assess sample quality and integrity. Samples that contain degraded material and/or DNA contaminants, which could affect library preparation performance, are flagged. Samples with a RIN below 2.5 are dropped from the study. Libraries were prepared using KAPA Stranded RNA-Seq Kit with RiboErase and unique Illumina-compatible PCR primers with indexes purchased from BioScientific (NEXTflex RNA-seq Barcodes, cat# 512915, 8nt index). Libraries were prepared using 500ng of total RNA input. Picogreen or a Qubit™ 2.0 Fluorometer were used to measure the total amount of RNA in the prepared library. Size distribution profiles of the final libraries are assessed using the Fragment Analyzer or Bioanalyzer. Libraries that fell outside of the expected size range and/or contain adapter dimer contaminants were flagged. Sequencing was performed either on an Illumina HiSeq 2500 sequencer (v4 chemistry) using 2x125bp cycles or on an Illumina NovaSeq 6000 sequencer using 2x100bp cycles. 40 million reads per sample were targeted.

### RNA-seq pre-processing

Samples were uniformly processed using RAPiD-nf, an efficient RNA-Seq processing pipeline implemented in the NextFlow framework^94^. Following adapter trimming with Trimmomatic (0.36)^95^, all samples were aligned to the GRCh38.primary_assembly build of the human reference genome using STAR (2.7.2a)^96^, with indexes created from GENCODE, version 30^97^. Gene expression was quantified using RSEM (1.3.1)^98^. Quality control was performed using SAMtools^99^ and Picard, and the results were collated using MultiQC^100^.

### RNA covariate selection

Model selection incorporated integrated quality control metrics, clinical data, and sample metadata to tailor covariate selection for individual tissues in a stepwise manner. For each tissue, linear models were fitted to each gene in the dataset to assess the individual effects of covariates. These models were ranked using the Bayesian Information Criterion (BIC) to prioritize those with the lowest BIC scores. A correlation analysis was then performed on the top-ranked models to eliminate highly correlated covariates, retaining only one candidate covariate for models exhibiting Pearson correlation scores greater than 0.9.

Variance partitioning was subsequently applied to the selected covariates to quantify their contributions to observed variations in gene expression. This was performed using variancePartition^101^ (1.30.2) in R. The top covariates contributing the most variance were then combined and evaluated using linear models and BIC scores. The covariate combination displaying the highest number of genes with the lowest BIC score was selected for subsequent analyses. This integrated approach ensured robust covariate selection across multiple tissues.

The resulting covariates, as shown below, were used in differential gene expression and deconvolution analyses. For analyses involving only ALS samples, the covariates were limited to library preparation type (automated KAPA or manual KAPA), sex, age at disease onset, and age at death. However, age at disease onset and age at death were excluded when computing differential expression for disease duration, age of onset, and age at death to avoid confounding effects. Library preparation for all hippocampus-derived samples was performed using an automated KAPA kit, and therefore, prep information was excluded from all analyses involving these samples.

### Differential expression

For all samples, except those derived from the motor cortex, differential expression (DE) analysis was performed using DESeq2^102^ (version 1.40.2) to compare gene expression patterns between ALS cases and non-ALS controls across individual tissue samples. DE analysis was also conducted on ALS samples to examine gene expression differences across five clinical features: *C9orf72* mutation status (present vs. absent), site of symptom onset (bulbar vs. limb), age of symptom onset, age at death, and disease duration. Age of symptom onset, age at death, and disease duration were treated as continuous variables. The raw count matrices served as the input for DE analysis, and curated covariates specific to each tissue type were included in the design formula. All other DESeq2 parameters were left at their default settings. For samples derived from the motor cortex, tissues were collected from either the lateral motor cortex, medial motor cortex, or an unspecified motor cortex region. For DE and downstream analyses, all motor cortex-derived samples were treated as originating from a single motor cortex region. Samples from the unspecified motor cortex region were obtained from single donors. However, some donors had samples from both the medial and lateral motor cortex regions. To account for multiple motor cortex samples originating from the same donors, DREAM^103^ was used for differential gene expression analysis through the variancePartition library (v1.30.2)^101^. The specific motor cortex region (medial, lateral, or unspecified) and donor IDs were included as random covariates.Gene set enrichment analysis and pathway analysis

Gene set enrichment analysis (GSEA)^104^ and general enrichment analysis were performed using the clusterProfiler R package^105^. For general enrichment analysis, the functions enrichGO (restricted to the Biological Process ontology) and enrichKEGG were used. Only significant differentially expressed genes (adjusted p-value < 0.05) were included in this step to identify enriched pathways and assess gene sharing across tissues. Cell marker GSEA was performed using the clusterProfiler package^105^ (version 4.8.3) to compare cell-type composition between ALS and control samples across tissue types. The input consisted of genes from the differential gene expression analysis, ranked by their log fold change (LFC) values. The reference cell marker gene sets were sourced from Kelley et al.^106^, PanglaoDB^107^, Mathys et al.^47^, NeuroExpresso^108^, and Darmanis et al.^109^.

### Signal sharing

Multivariate Adaptive Shrinkage (MASH)^45^ was used to assess signal sharing and similarity in differential gene expression between tissues, utilizing the mashr v0.2.79 library in R. The log_2_ fold-change values, along with their standard errors from differential expression (DE) analysis, were used as input. Any gene with an absolute log_2_ fold-change value greater than 5 in at least one tissue was excluded to minimize the impact of outliers. Signal sharing was specifically calculated using the `get_pairwise_sharing` function with default settings.

### Cell-type deconvolution

Cell-type deconvolution was performed using MuSiC^110^ (1.0.0). For cerebellum-derived samples, the GSE246183 single-cell dataset served as the reference^49^. The GSE190442 spinal cord–derived reference dataset was used for spinal cord samples^48^. For the remaining samples, a cortex-derived single-cell dataset (GSE140231) was used as the reference^47^. For each tissue reference, only donors corresponding to non-neurological disease controls were used. To account for potential confounding effects of covariates, curated covariates for each tissue were used when assessing differences in cell-type composition between ALS and control samples across tissues. This was achieved by fitting a linear model to the computed cell-type composition, using the selected covariates as predictors, and then adjusting the cell-type composition based on the residuals of the fitted model.

### Differential Splicing

Splice junctions were extracted from the STAR-aligned RNA-seq bam files with regtools^111^ (0.5.2) ‘junctions extract’ using default parameters. Junctions from all samples were clustered together using the Leafcutter ^51^ (v2c9907e) script `leafcutter_cluster_regtools.py` to allow comparison of DS clusters between tissues and analyses. For differential splicing analysis between binary outcomes, leafcutter was run with ‘-t 600 -p8’ parameters, with covariates selected per tissue as for the DE analysis above. For DS analyses on continuous traits the leafcutter2 pipeline was used with default parameters. Introns were annotated using the leafidex pipeline. Splicegraph visualizations were constructed using the leafviz pipeline (see **URLs**). All DS statistical tests utilized a Dirichlet-multinomial generalized linear model and were corrected for multiple comparisons when necessary via the Benjamini–Hochberg procedure.

For gene ontology enrichment analysis, for each tissue and comparison the background set was set to the tested genes for that analysis, that passed QC for DE analysis as above. DS genes were determined from introns using Leafidex pipeline output. We used the “enrichGO”, “enrichKEGG”, (clusterProfiler v3.0.4) and “enrichPathway” (ReactomePA v.16.2) functions to query the GO BP, KEGG, and Reactome DBs respectively, and filtered results to FDR <0.05. To assign DS clusters to potentially contributing cell types we utilized the single cell references described above for the relevant tissues^47–49^. Summary statistics produced by the Seurat “FindMarkers” function were subset to those with avg_log2FC > 0 to select for those enriched in particular cell types. Z-scores and avg_log2FC were multiplied to combine a composite ‘score’ for each gene and cell type, and the highest score was taken for each gene to assign DS clusters to potentially contributing cell types.

### Quantitative Trait Locus Mapping

To perform expression QTL (eQTL) mapping, we used a previously published pipeline based on the one created by the GTEX consortium^10^. Gene expression matrices were created from the RSEM outputs using tximport^112^. Matrices were then converted to GCT format, TMM normalized, filtered for lowly expressed genes, removing any gene with less than 0.1 TPM in 20% of samples and at least 6 counts in 20% of samples. Each gene was then inverse-normal transformed across samples. PEER factors were calculated to estimate hidden confounders within our expression data. 10 PEER factors were estimated for tissues with sample size less than 250; and 15 PEER factors for tissues with more than 250 samples. We created a combined covariate matrix that included the PEER factors, the first 5 genotyping principal component values, sex, age, DNA and RNA library preparation as input to the analysis.

To test for cis-eQTLs, linear regression was performed using the tensorQTL^113^ *cis_nominal* mode for each SNP-gene pair using a 1 megabase window within the transcription start site (TSS) of a gene. To test for association between gene expression and the top variant in cis we used tensorQTL cis permutation pass per gene with 1000 permutations. To identify eGenes, we performed q-value correction ^114^ of the permutation P-values for the top association per gene at a threshold of 0.05.

We performed splicing quantitative trait loci (sQTL) analysis using the splice junction read counts generated by regtools (v0.5.1)^111^. Junctions were clustered using Leafcutter ^51^, specifying for each junction in a cluster a maximum length of 500,000 bp. Following the GTEx pipeline, introns without read counts in at least 50% of samples or with fewer than 10 read counts in at least 10% of samples were removed. Introns with insufficient variability across samples were removed (min usage ratio 1%). Junctions missing in more than 50% of samples in every tissue-cohort pair were removed, along with those with PSI values of 0 or 1 in 90% of samples. Filtered counts were then scaled, centered and normalized using `prepare_phenotype_table.py` from Leafcutter, merged, and converted to BED format, using the coordinates from the middle of the intron cluster. We created a combined covariate matrix that included 15 PEER factors and the first 5 genotype principal components, the first 5 genotyping principal component values, sex, age and DNA and RNA library preparation as input to the analysis.

To test for cis sQTLs, linear regression was performed using the tensorQTL nominal pass for each SNP-junction pair using a 1Mb window from the center of each intron cluster. To test for association between intronic ratio and the top variant in cis we used tensorQTL permutation pass, grouping junctions by their cluster using --grp option. To identify significant clusters, we performed q-value^114^ correction using a threshold of 0.05. We estimated pairwise replication (π_1_) of eQTLs and sQTLs using the q-value R package (2.28.0). This takes the SNP-gene pairs that are significant at q-value < 0.05 in the discovery dataset and extracts the unadjusted P-values for the matched SNP-gene pairs in the replication dataset.

### GWAS integration

We acquired full summary statistics from the latest ALS GWAS^1^, using the ALL meta-analysis of European and East Asian populations. Genome-wide significant loci were taken to be the most significant variants within 1 megabase at a threshold of P < 5e-8, removing loci overlapping the major histocompatibility complex region on chromosome 6. We used COLOC (v5.2.3)^115^ to test whether SNPs from each locus in the GWAS colocalized with expression and splicing QTLs. For each locus in the ALS GWAS we extracted the nominal summary statistics of association for all SNPs within 1 megabase either upstream/downstream of the top lead SNP (2Mb-wide region total). In each QTL dataset we then extracted all nominal associations for all SNP-gene pairs within that range and tested for colocalization between the GWAS locus and each gene. We set a threshold for colocalization as posterior probability of colocalization, hypothesis 4 (PP4) value > 0.9. To avoid spurious colocalization caused by long range linkage disequilibrium, we restricted our colocalizations to GWAS SNP - eQTL SNP pairs where the distance between their respective top SNPs was ≤ 500kb or the two lead SNPs were in moderate linkage disequilibrium (R^2^ > 0.1), taken from the 1000 Genomes (Phase 3) European populations using the LDLinkR package. For splicing QTLs we followed the same approach but collapsed junctions to return only the highest PP4 for each gene in each locus. We restricted reported sQTL colocalizations to cases where the GWAS SNP and the top sQTL SNP were either within 100kb of each other or in moderate linkage disequilibrium (R^2^ > 0.1).

### STR QTLs

After merging single VCFs into a multi-sample VCF, we performed both call-level and locus-level filtering using dumpSTR from the TRTools toolkit (v4.0.1) ^81^. The filtering was conducted with the following parameters: `--min-locus-hwep 0.000001 --filter-regions hg38_segdup.sorted.bed.gz --min-locus-callrate 0.4 --filter-regions-names SEGDUP`. dumpSTR removed TRs with low call rate, TRs whose genotypes do not follow Hardy-Weinberg Equilibrium and TRs overlapping segmental duplications downloaded from the UCSC Genome Browser. We additionally used the parameter --eh-min-call-LC 10 to remove low quality calls for ExpansionHunter. 47 out of 68 tandem repeats passed these filters.

We analyzed 456 European samples with PCR-free WGS data and matched RNA-seq data for QTL mapping: 223 in Cerebellum, 301 in Cervical_Spinal_Cord, 329 in Frontal_Cortex, 70 in Hippocampus, 115 in Lateral_Motor_Cortex, 285 in Lumbar_Spinal_Cord, 108 in Medial_Motor_Cortex, 52 in Occipital_Cortex, 96 in Temporal_Cortex, 43 in Thoracic_Spinal_Cord, 163 in Motor_Cortex_BA4unspecified. The ExpansionHunter output has two estimated size values corresponding to the two alleles for each TR. Each value represents the number of motifs present on one allele. We refer to the shorter one as *min_allele* and the longer one as *max_allele*. Only TRs with a *max_allele* variance of at least 1 across samples within each tissue were included in the analysis. To define the thresholds for grouping, we set the first threshold (t_1_) as the mean of *min_allele* across samples within each tissue, assuming it represented the unexpanded state. The second threshold (t_2_) was defined as t_1_ plus the standard deviation of *max_allele* across samples within each tissue. Samples with a TR size ≤ t_1_ were classified as small, those between t_1_ and t_2_ as medium, and those ≥ t_2_ as large. The groups were encoded as 0 (small), 1 (medium), and 2 (large).

We used tensorQTL (v1.0.10) in map_cis mode to identify associations between tandem repeat (TR) sizes and gene expression and splicing junction usage. Analyses were conducted separately for each tissue, with the top four principal components (PCs) from SNP genotype data (scikit-learn v1.3.1) included as covariates. The sequencing platform used for WGS preparation (HiSeq, NovaSeq V1, or NovaSeq V1.5) was also included as a covariate after conversion to dummy variables. Gene expression and junction matrices were the same as used for standard QTL mapping, then residualised for the same number of PEER factors as QTL mapping. We limited our analysis to genes and splice junctions within 1 megabase of each TR. To evaluate statistical significance, tensorQTL uses a permutation-based approach to generate an empirical null distribution for each phenotype. Specifically, we used the default setting of 10,000 permutations to estimate permutation-based p-values. These p-values were then corrected for multiple testing using the Benjamini–Hochberg procedure. Multiple testing correction was performed separately for each type of molecular phenotype (gene expression or splicing) within each tissue.

### Allele-specific expression

Allele-specific expression (ASE) analysis was performed using phASER (v1.2.0)^116^ using donors with paired WGS and RNA-seq data in at least one tissue. Briefly, VCF files were phased using SHAPEIT4 (v4.1.3)^117^ using the 1000 Genomes reference panel^75^ The *phaser.py* script was run on all of the RNA-seq samples for each VCF to identify haplotypes. The *phaser_ae.py* script was used to aggregate the counts at each gene using GENCODE v25. The aneva-h tool (v0.0.0.9000)^58^ was used to estimate the variance in dosage from genetic variation (VG) for each gene in each tissue. Finally, ANEVA-DOT (v0.1.1) was used on each individual-tissue combination with the appropriate tissue VG estimates to identify ASE outliers in each individual for each tissue^58^.

π_1_ estimates were calculated to determine the degree of individual-outlier tissue sharing using the qvalue package (v2.38.0)^59^. This approach estimates the proportion to true positive ASE outliers in tissue *j* using the p-value distribution of ASE outliers in tissue *i*. The median π_1_ of each tissue-tissue comparison across the entire cohort was visualized as a heatmap with dendrograms showing hierarchical clustering from the ComplexHeatmap package (v2.22.0)^118^.

A meta-analysis was performed by using Fisher’s method to combine p-values across all of the tissues for each individual with more than one tissue’s RNA-seq. To determine the enrichment of ASE outliers in ALS cases vs controls, or between individuals that do or do not have *C9orf72* repeat expansion, we performed one-sided Fisher’s exact tests for each gene. The neuronal Hi-C data and TAD calls from Rahman et al^61^ were used to visualize the 3D chromatin structure at the *C9orf72* locus using the plotgardener package (v1.12.0)^119^.

To annotate rare variants (RVs), variant annotations from the joint VCF across the entire cohort (method described above). RVs affecting a gene were defined as being located within the gene body and ± 10kb of the gene body and having a MAF < 0.01 from 1000 Genomes^75^. If there were multiple variants for each individual-gene, one variant was selected according to the following hierarchy: large deletion, large insertion, splice acceptor, splice donor, splice region, frameshift, stop, TSS, conserved noncoding, coding, other noncoding, no variant. Large insertions were further categorized into retrotransposable elements (Alu, SVA, LINE1). Since the number of non-ASE outlier individual-genes far exceeded the number of ASE outlier individual-genes, each category was normalized according to the total number of variants belonging to each category (ASE outlier, non-ASE outlier). Then within each category, a proportion of ASE outliers to non-ASE outliers was calculated.

## Data Availability

All supplementary tables, including donor and sample-level metadata, along with gene expression and splicing count data, QTL and rare variant summary statistics have been uploaded to Zenodo: https://zenodo.org/communities/nygc_als

All WGS data has been deposited through dbGAP (accession phs003067.V3) to AnViL. 4,851 WGS samples, including 107 additional samples not analyzed in this manuscript, reside in 3 data buckets according to access restrictions: general research use (n=1,964), health/medical/biomedical (n=2,768), disease-specific (genetic diseases of muscle, nerve, spinal cord or brain; n=12) (see **Table S1**). Two out of 4,746 analyzed samples are unavailable for download due to sharing restrictions. The integrated 4,744-sample joint VCF including SNV, INDEL, and SV calls (both full callset and stringent QC subset) as well as a site-level VCF with VEP annotations can be found in the disease-specific bucket. WGS data will be available on the AnViL platform from August 2026. Until then, access can be gained by emailing https://cgnd_help@nygenome.org.

All RNA-seq data (FASTQs, gene counts table) has been deposited on the gene expression omnibus (GEO; accession GSE137810). This comprises data on 2,906 samples, including 332 additional samples not analyzed in this manuscript. Additionally, RNA-seq data are being uploaded to the ALS Knowledge Portal (https://ampals.synapse.org/), at the following accessions: syn67729513, syn67733559, syn67737779.

Finally, we have created an interactive website to explore all gene expression associations across the five largest tissues, available at https://alsbrowser.nygenome.org/

## Code Availability

All code written for the project has been deposited on GitHub: https://github.com/jackhump/NYGC_ALS_Flagship

## URLs

Absinthe: https://github.com/nygenome/absinthe

EH v5 variant catalog: https://github.com/nygenome/nygc-germline-pipeline-readmes/blob/master/EHv5_variant_catalog.json.

GIAB low complexity genomic stratification BED file: https://ftp-trace.ncbi.nlm.nih.gov/ReferenceSamples/giab/release/genome-stratifications/v3.3/GRCh38@all/LowComplexity/GRCh38_AllTandemRepeatsandHomopolymers_slop5.bed.gz

leafcutter2: https://github.com/davidaknowles/leafcutter/tree/leafcutter2

leafidex: https://github.com/atokolyi/leafidex

leafviz: https://github.com/jackhump/leafviz

PicardTools: https://broadinstitute.github.io/picard

## Author Contributions

Conceptualization, J.H., G.N., D.F., G.G., S.F., and H.P.; data curation, J.H., A.O.B., M.B.B., A.C., A.O., A.T., K.B., U.S.E., H.G., B.J., W.H.D., B.N.H., A.Ru., F.P.S., G.N., D.F., and S.F.; formal analysis, J.H., A.O.B., M.B.B., A.C., A.O., A.T., K.B., A.R., Y.K., M.L.B., U.S.E., W.E.C., H.G., S.C., R.A., D.F., G.G., and M.B.H.; investigation, J.H., A.O.B., M.B.B., A.C., A.O., A.T., U.S.E., W.E.C., H.G., D.M., J.Mc., G.N., N.R., D.F., G.G., and M.B.H.; methodology, J.H., A.O.B., M.B.B., A.C., A.O., A.T., K.B., A.R., U.S.E., W.E.C., R.F., H.G., T.N., D.M., J.Mc., N.P., G.N., N.R., D.F., and G.G.; software, J.H., A.O.B., M.B.B., A.C., A.O., A.T., K.B., A.R., U.S.E., W.E.C., R.F., H.G., T.N., R.M., W.H.D., and G.G.; validation, A.O.B., M.B.B., A.C., A.O., A.T., U.S.E., W.E.C., and H.G.; visualization, J.H., A.O.B., M.B.B., A.C., A.O., A.T., U.S.E., H.G., and G.N.; project administration, J.H., M.H.P., A.Ru., N.P., S.F., D.F., and H.P.; resources, T.R. and M.B.H.; supervision, J.H., M.C.Z., H.H.W., G.N., N.R., T.L., D.A.K., T.R., and H.P.; funding acquisition, S.F., G.G., and H.P.; writing - original draft preparation, J.H., A.O.B., M.B.B., A.C., A.O., A.T., A.R., M.L.B., W.E.C., A.Ru., S.F., G.N., N.R., D.F., and G.G.; writing - review & editing, all authors.

## Acknowledgments

We thank the patients and families who donated material. This study was supported by the following National Institutes of Health grant #s: NIA U01-AG058635, NIA R21-AG063130, NIA R01-AG054005, NIA U01-AG068880, NIA RF1-AG065926, NIA R56-AG055824, NIA P30-AG066514, NINDS U54-NS123743, NINDS R01-NS116006, NIH OD R03-OD036491, NIH U19AG074862, NIA U54AG075932, NIA U54AG076040, NIA R01AG066831, NIA R01NS118570, NINDS R01NS118183, NINDS R01NS127186, NINDS R01NS117583, NINDS R01NS116350, NINDS OT2NS136939 and NINDS RF1NS139948 to T.R., J.H., B.J., K.BP, W.H.D., T.N., G.G. and H.P. J.H. is supported by Target ALS, My Name’5 Doddie, the Packard Center for ALS Research, and the BrightFocus foundation. H.P. is additionally supported by Target ALS grants BB-2022-C7-L2, FD-2023-GEN-S1, CF-2023-GEN-S1 and NHGRI RM1HG011014, the ALS Association, the Tow Foundation, Mitsubishi Tanabe grant, and the New York State grant. This work was supported in part through the computational resources and staff expertise provided by Scientific Computing at the Icahn School of Medicine at Mount Sinai and supported by the Clinical and Translational Science Awards (CTSA) grant no. UL1TR004419 from the National Center for Advancing Translational Sciences. Research reported in this paper was supported by the Office of Research Infrastructure of the National Institutes of Health under award numbers S10OD026880 and S10OD030463. The content is solely the responsibility of the authors and does not necessarily represent the official views of the National Institutes of Health.

## Supplementary Tables - hosted on Zenodo

https://zenodo.org/communities/nygc_als

All tables can be read directly into R using the readxl package:

`df <- readxl::read_excel(path = “path_to_file”, sheet = “ST1”, skip = 1)`

## Metadata

Dictionary: all column descriptions in tables S1-S3

ST1: all WGS and donor-level clinical metadata

ST2: WGS mutation annotations

ALS genes: All ALS genes tested in WGS

ST3: all RNA-seq metadata

## Expression tables

ST4: all case-control DEGs across tissues

ST5: all GSEA GO analysis merged, with column for comparison and tissue

ST6: all GO enrichment for tissue intersections

ST7: all deconvolution estimates across all tissues and cell-types

ST8: all deconvolution statistics, across all tissues and cell-types

ST9: cell-type marker GSEA results

ST11-15: clinical DEGs

ST16: clinical GO/KEGG enrichment on age death and duration

## Splicing tables

ST17-22: Categorical Case-control, C9, limb-bulbar DS results at FDR < 0.05 (clusters and introns)

ST23-28: Continuous DS results, onset, death, duration (clusters and introns)

ST29-31: DS enrichments

## Genetics tables

ST32: QTL discovery

ST33: QTL sharing

ST34: COLOC results

ST35: STR-QTL results

## Supplementary Data - hosted on Zenodo

https://zenodo.org/communities/nygc_als

Gene expression counts and transcripts per million values

Splice junction counts

eQTL summary statistics

sQTL summary statistics

Allele-specific expression counts and outlier scores

## Supplementary Figures

**Figure S1:** SNV/INDEL Calls

**Figure S2:** SV calls

**Figure S3:** EH STRs

**Figure S4:** Mutations in FTD and other MND

**Figure S5:** RNA-seq QC - variancePartition plots for the 5 main tissues and PCA

**Figure S6:** DEG discovery per tissue against sample size

**Figure S7:** GSEA with markers

**Figure S8:** Clinical Upset plots, GO and KEGG

**Figure S9:** Deconvolution in controls

**Figure S10:** Additional splicing results

**Figure S11:** QTL sharing

**Figure S12:** Full STR-QTL results

**Figure S13:** Additional characterization of ASE outliers

## Methods tables

Methods table 1 - library preparation types

Methods table 2 - variant discovery

Methods table 3 - covariates used for RNA-seq

## Supplementary Figures

**Figure S1.**
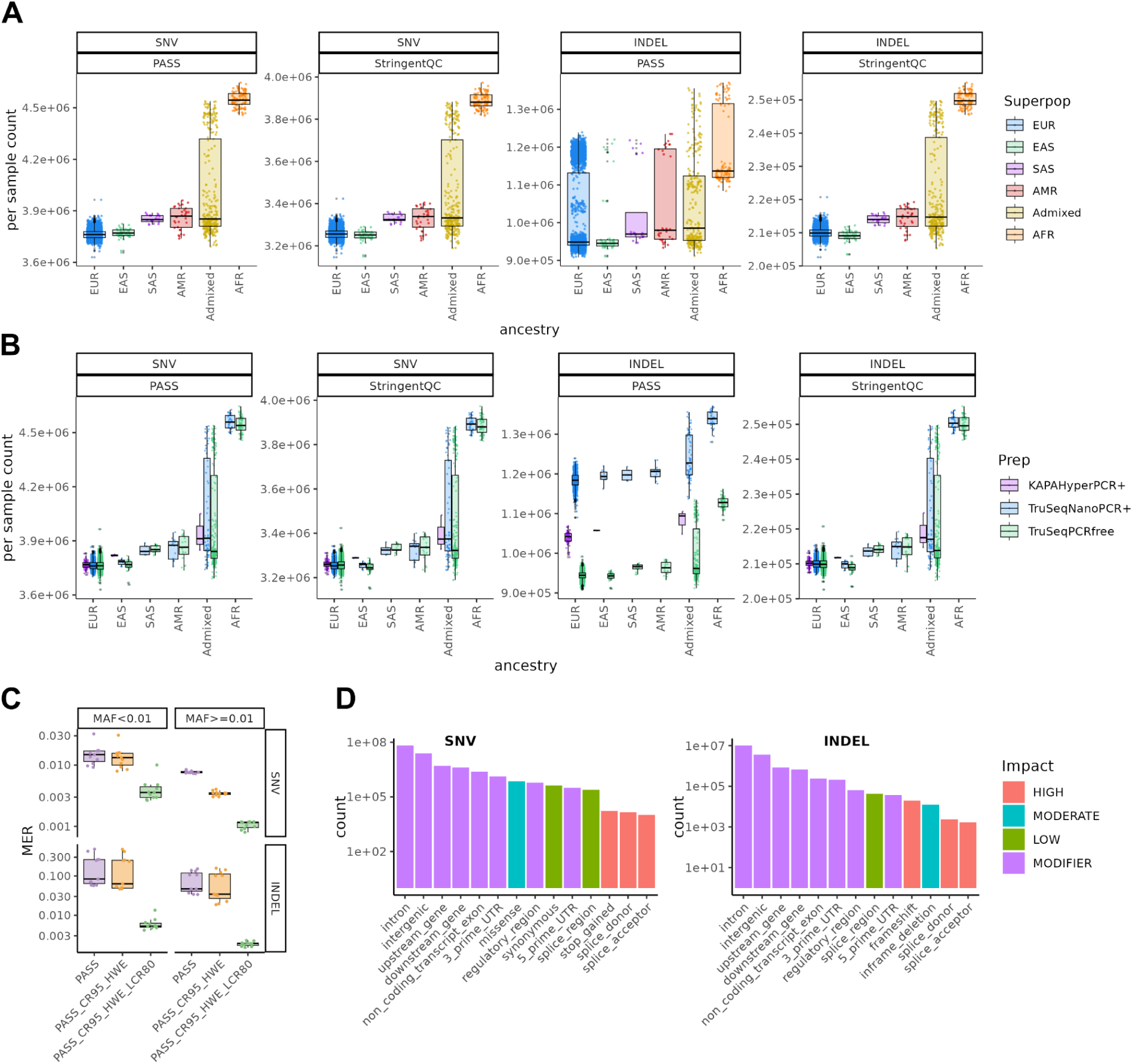
| Small variant calls in WGS data across 4,746 samples. **A)** Per sample small variant count distributions stratified by ancestry group (EUR: European, EAS: East Asian, SAS: South Asian, AMR: American, AFR: African) and filtering condition, and by **B)** library prep kit type. **C)** Mendelian error rate (MER) of called genotypes stratified by variant type, minor allele frequency (MAF), and three filtering conditions (from left to right: VQSR PASS, intermediate, StringentQC) computed for 11 child samples in mother-father-child trios. CR95: genotype call rate > 95%, HWE: HWE exact test p-value > 1e-10, LCR80: low complexity region overlap < 80%. **D)** Genomewide counts of Variant Effect Predictor (VEP) consequences (most severe) stratified by variant type (PASS sites). Fill color represents impact category (high, low, moderate, modifier), as defined by VEP.

**Figure S2.**
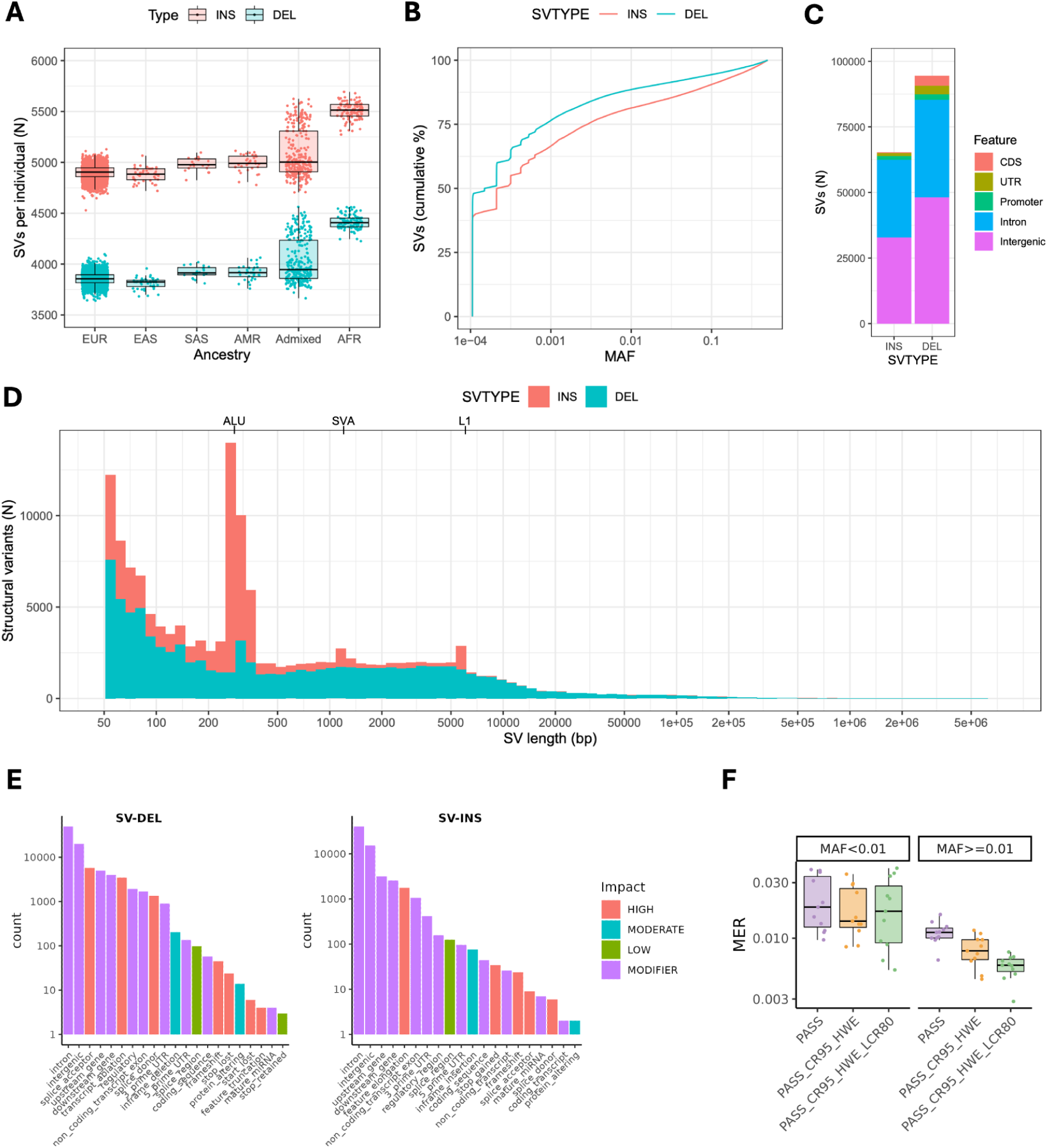
| Structural variant discovery from WGS data across the 4,746 samples. **A)** Per sample counts, stratified by ancestry group (EUR: European, EAS: East Asian, SAS: South Asian, AMR: American, AFR: African). The Admixed category represents individuals for which no individual ancestry component was above 80%. **B)** Cumulative distribution of minor allele frequency (MAF) for insertions and deletions. **C)** Stacked barplot representing SV overlap with genic features. For SVs overlapping multiple feature types, a single count, prioritized in the following order: CDS, UTR, Promoter, Intron and Intergenic, is reported. **D)** Stacked histogram representing the size distribution of insertions and deletions. **E)** Genomewide counts of Variant Effect Predictor (VEP) consequences (most severe) stratified by variant type (PASS sites). Fill color represents impact category (high, low, moderate, modifier), as defined by VEP. **F)** Mendelian Error Rate (MER) of called genotypes stratified by variant type, minor allele frequency (MAF), and three filtering conditions (from left to right: VQSR PASS, intermediate, StringentQC) computed for 11 child samples in mother-father-child trios. CR95: genotype call rate > 95%, HWE: HWE exact test p-value > 1e-10, LCR80: low complexity region overlap < 80%.

**Figure S3.**
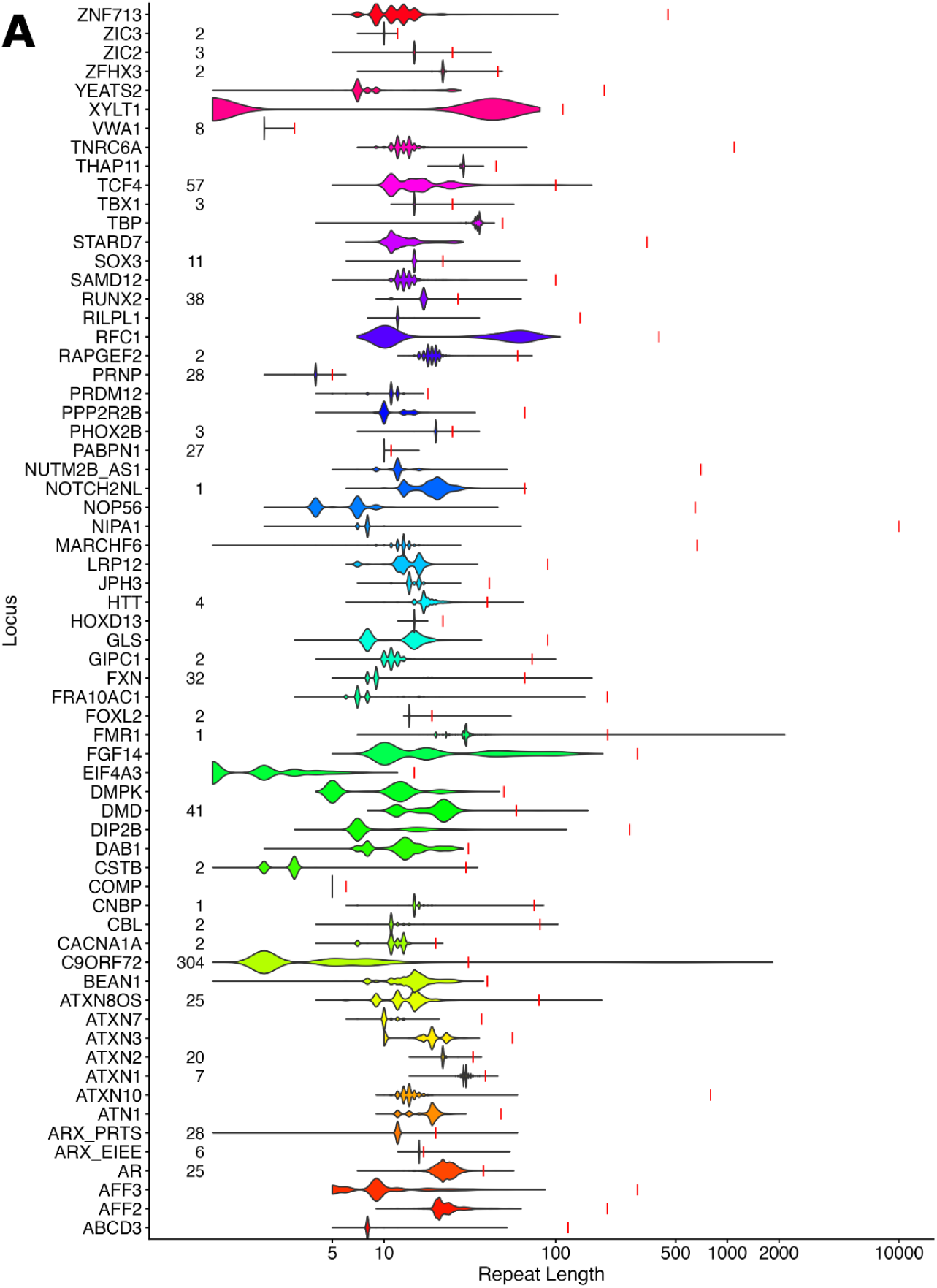
| Clinical STR discovery. (**A**) ExpansionHunter repeat length distributions of both alleles for 3,465 PCR-free samples genotyped across 68 loci. Vertical red lines represent the pathogenic minimum for each locus. The count of samples with a pathogenic expansion is listed to the left of the violin plot for each affected locus.

**Figure S4.**
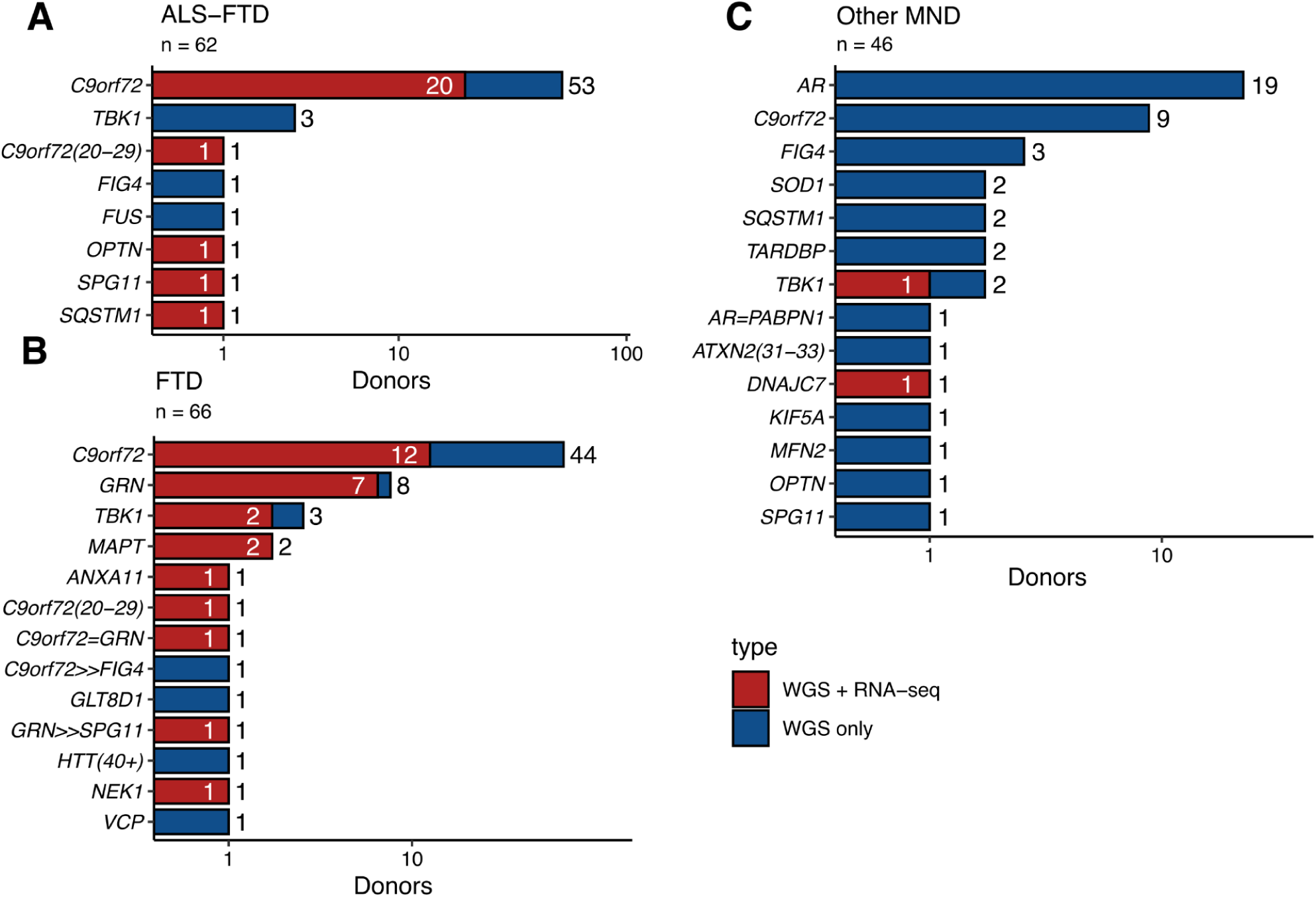
| Annotated mutations discovered in other diseases. Breakdown of known mutations discovered in ALS with frontotemporal dementia (ALS-FTD; **A)**, pure FTD (**B**), and other motor neuron diseases, including spinal bulbar muscular atrophy, primary lateral sclerosis, and progressive bulbar palsy (**C**). Patients with mutations in multiple genes denoted with a plus symbol.

**Figure S5.**
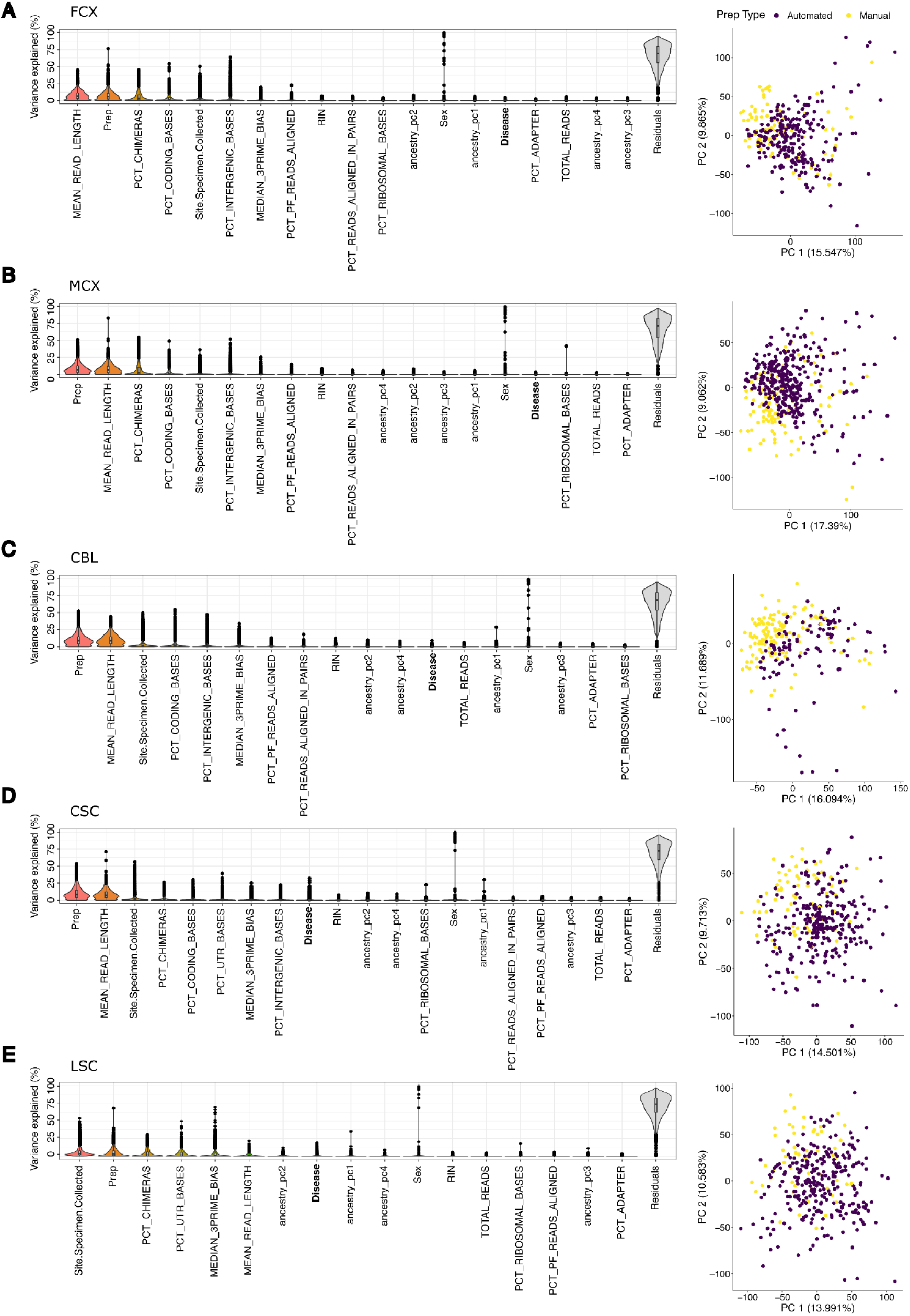
| RNA-seq quality control and covariate selection. Left panels: VariancePartition plots for each tissue (**A-E**). In each gene a linear mixed model is fitted to test the variance explained by a set of largely uncorrelated clinical and technical variables. Variables are ranked by the median variance explained across all genes. Library preparation method is among the top variables for all tissues. Variance explained by disease is highlighted in bold font. **Right panels**: principal components analysis plot of each tissue, coloured by library preparation method.

**Figure S6.**
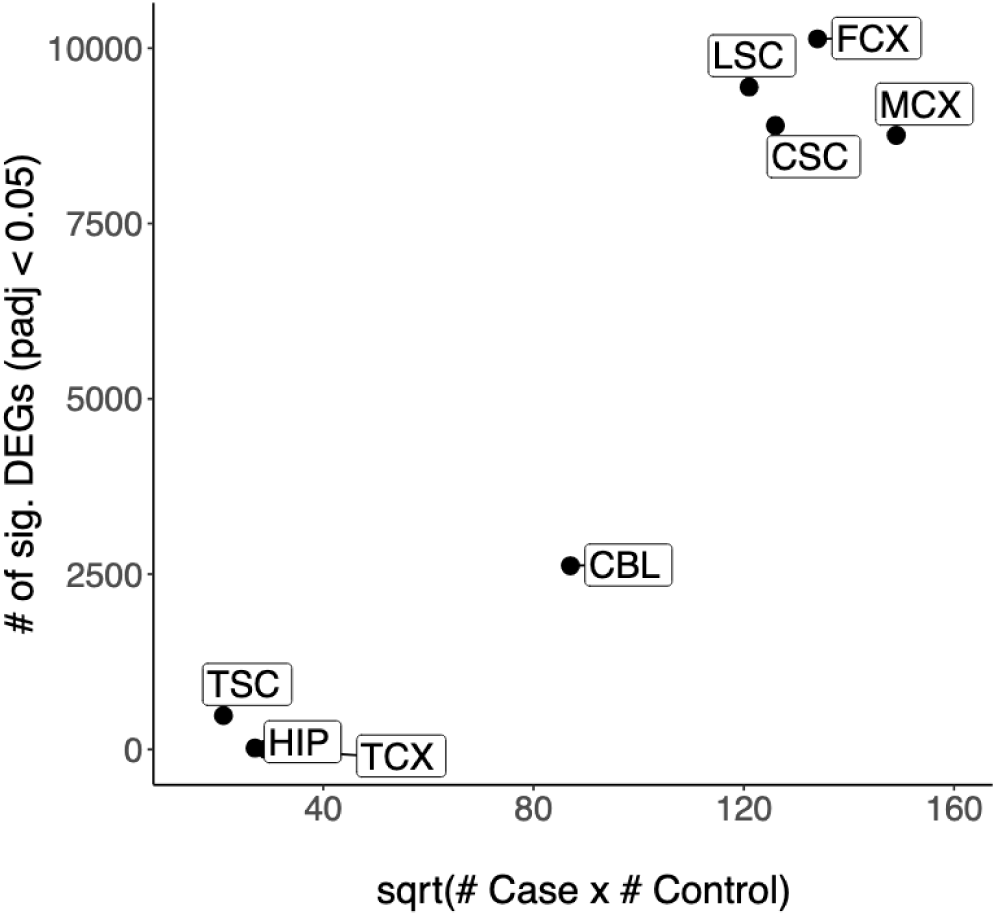
| Differential gene expression discovery against sample size. Sample size presented as the effective sample size, equal to the square root of the product of cases and controls.

**Figure S7.**
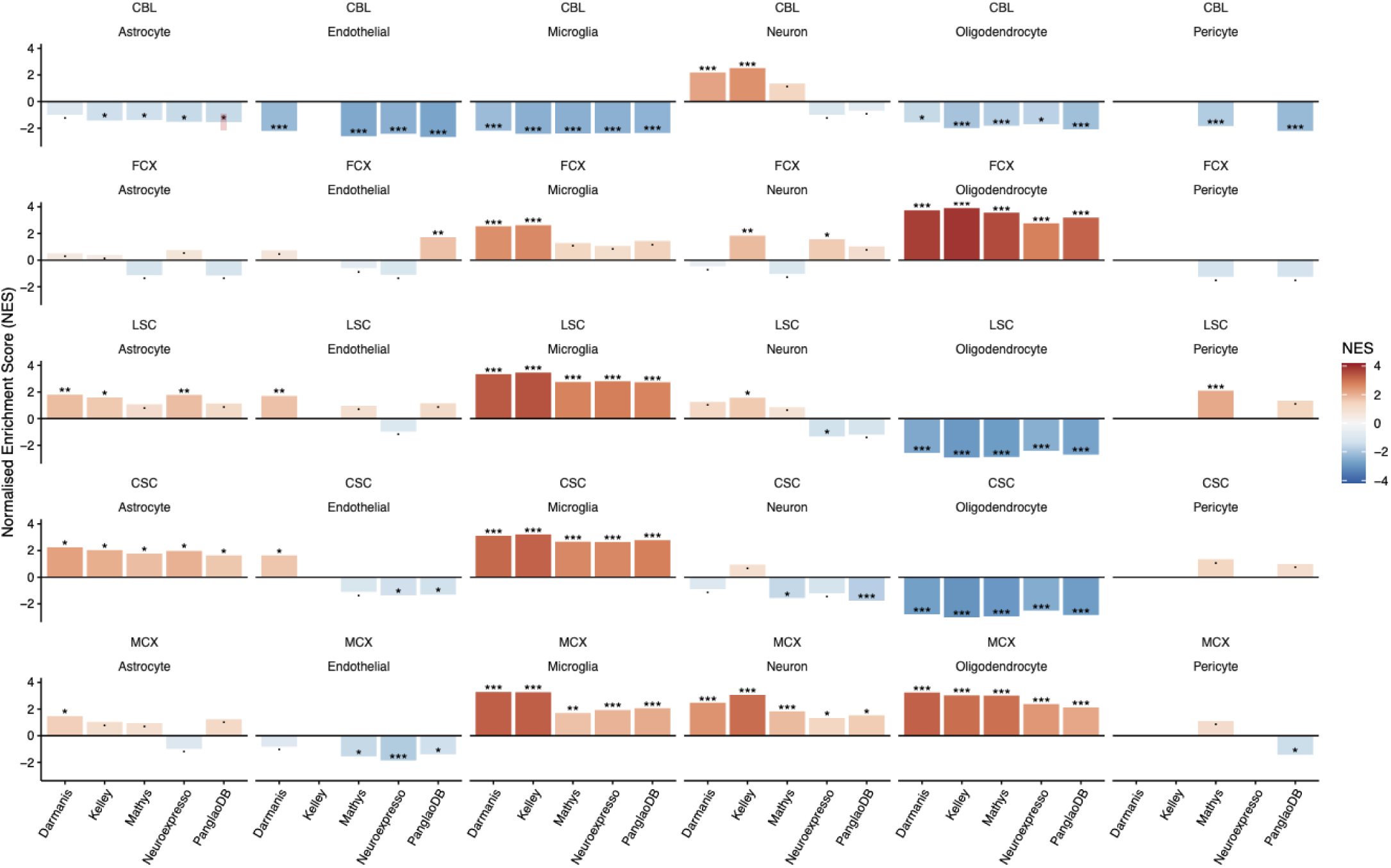
| Individual cell marker gene set enrichment analysis. Each row corresponds to a tissue, with each panel showing the normalized enrichment score (NES) from GSEA comparing the enrichment of cell-type markers from 5 different databases in ALS vs Control DEG summary statistics. Significance is indicated as follows: * padj < 0.05; ** padj < 0.001; *** padj < 0.0001.

**Figure S8.**
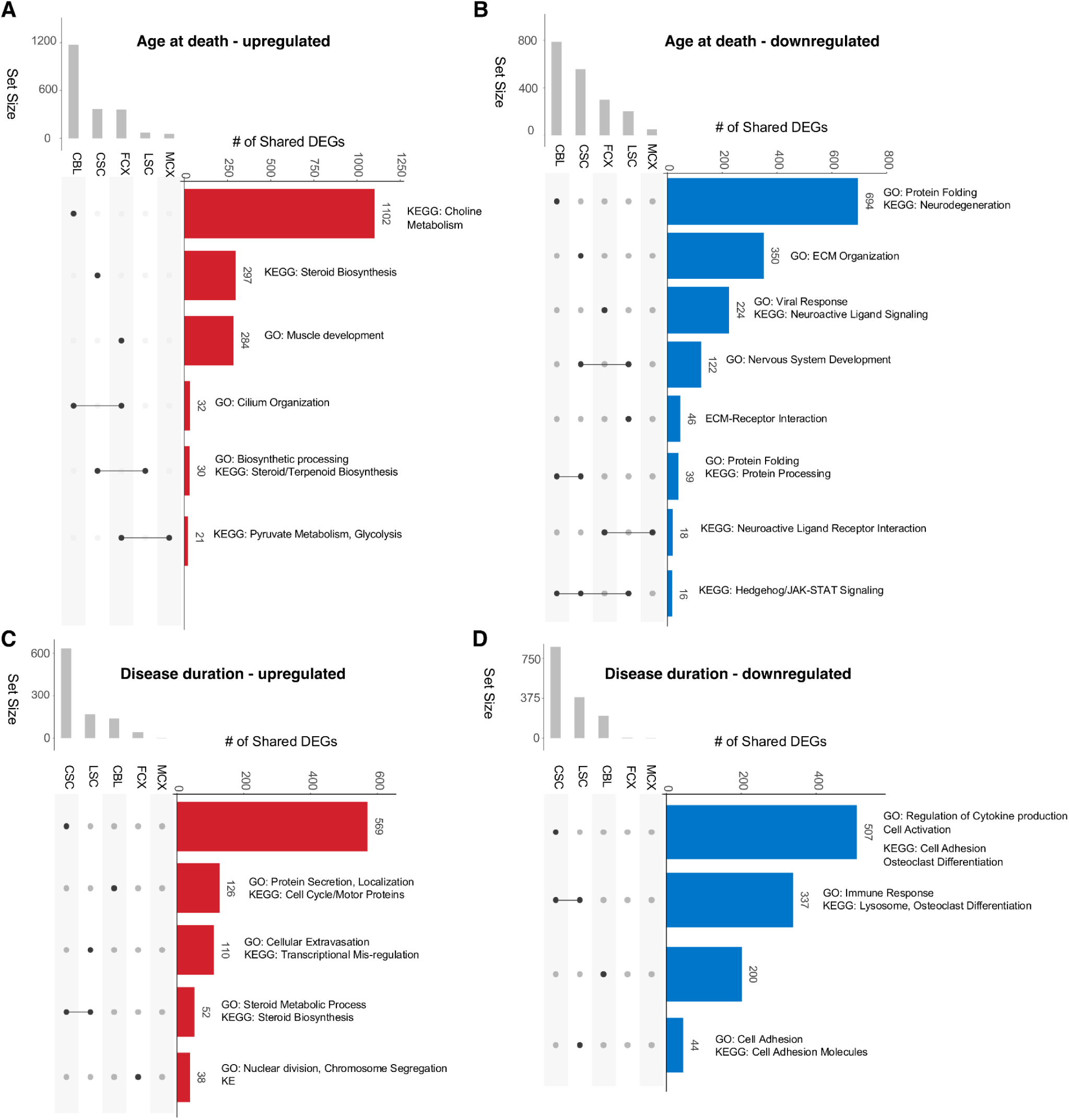
| Shared genes associated with age at death and disease duration. A-D) UpSet plots show intersecting gene sets across tissues with age at death (upper panels) and disease duration (lower panels), split into shared upregulated (left panels) and downregulated (right panels). Only the largest intersections are shown. Intersecting sets tested with GO and KEGG for enriched pathways. Top pathways shown adjacent to each bar.

**Figure S9.**
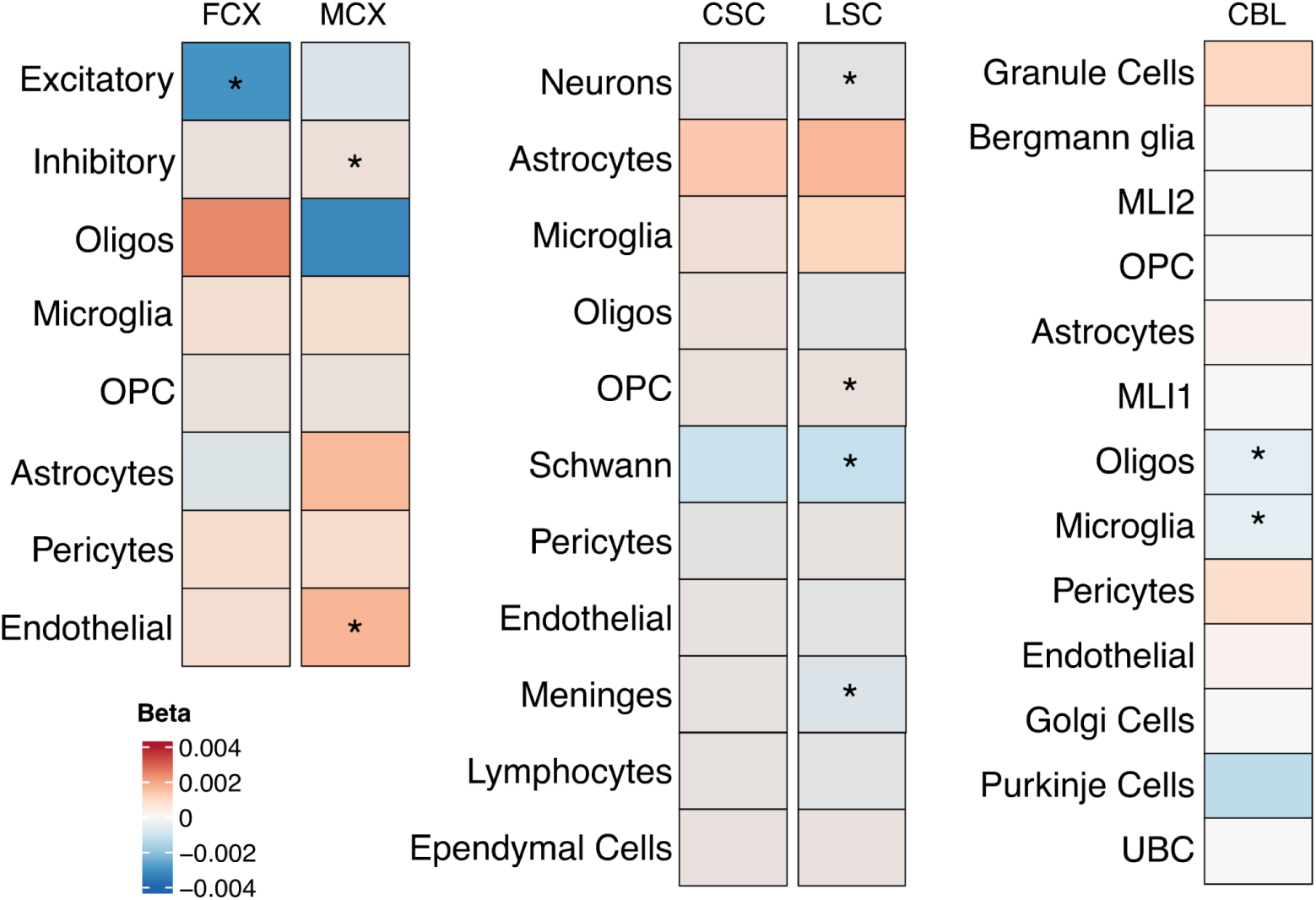
| Age of death correlations in control donors. Each association tested using a linear model adjusting for prep kit type and sex, with tissue origin included as a covariate for motor cortex only. P-values adjusted for number of cell-types tested. *: Bonferroni-adjusted P < 0.05.

**Figure S10.**
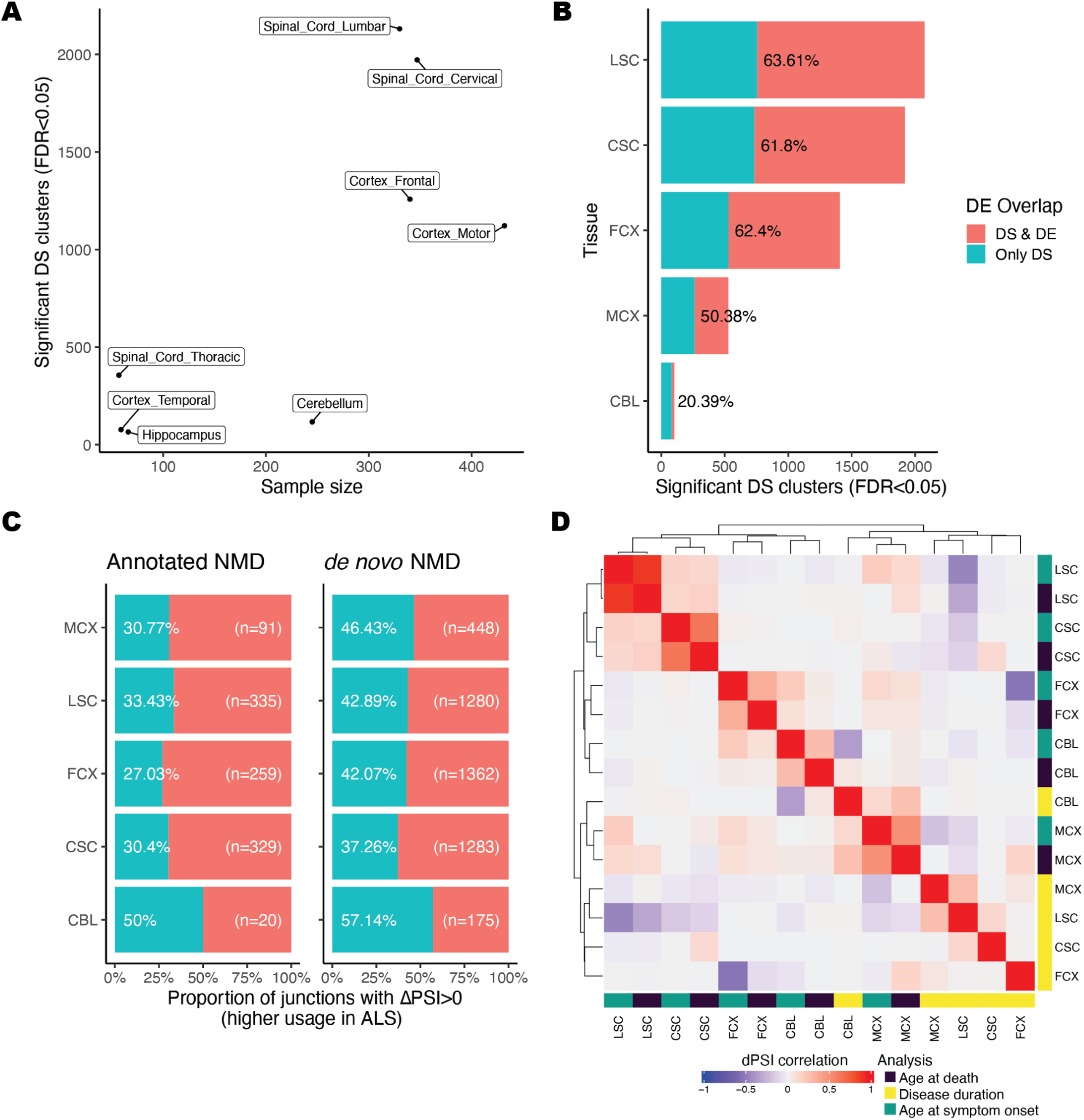
| Additional information for the differential splicing analysis. **A)** Scatter plot showing the relationship between tissue sample size (number of individuals, x axis), and the number of significant (FDR <0.05) differentially spliced clusters (y axis). **B)** Bar plot showing the count of differentially spliced clusters (FDR <0.05) that were within genes that were also differentially expressed (in red), or only differentially spliced (in blue). This is separated by tissue (y axis), with percentages indicating the proportion of DS clusters that were in differentially expressed genes. **C)** Bar plot showing the proportion of NMD junctions annotated (left), or *de novo* predicted (right), that have a ΔPSI>0 representing higher usage in ALS cases (blue, %). The number of NMD junctions per tissue is displayed on the right in white. **D)** Heatmap showing for each major tissue the Pearson correlation of ΔPSI scores for the age at death, age at symptom onset, and disease duration comparisons. X and y axes are hierarchically clustered.

**Figure S11.**
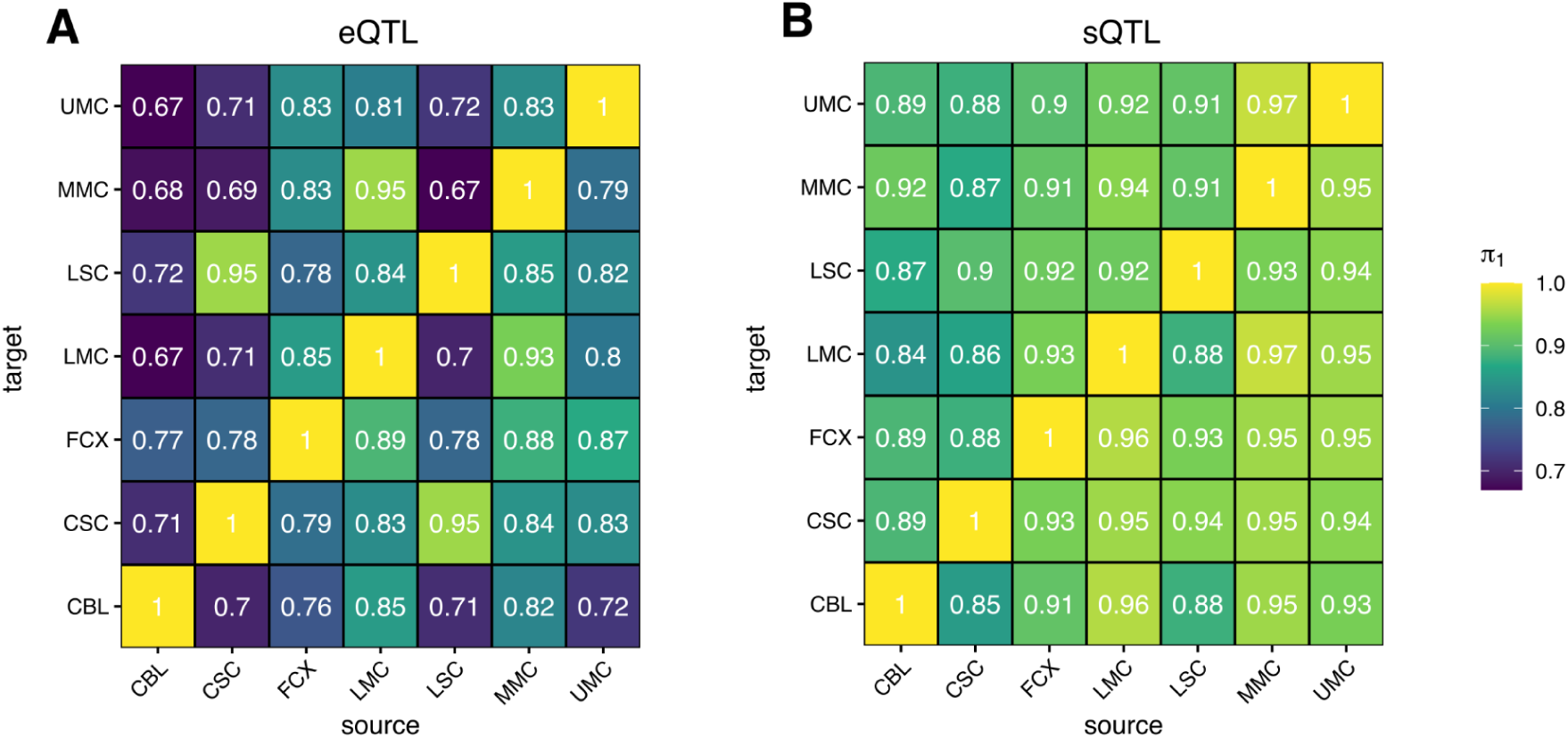
| Sharing of QTL effects across tissues. Pairwise sharing of genetic effects on eQTLs (**A**) and sQTLs (**B**) using Storey’s π_1_ metric.

**Figure S12.**
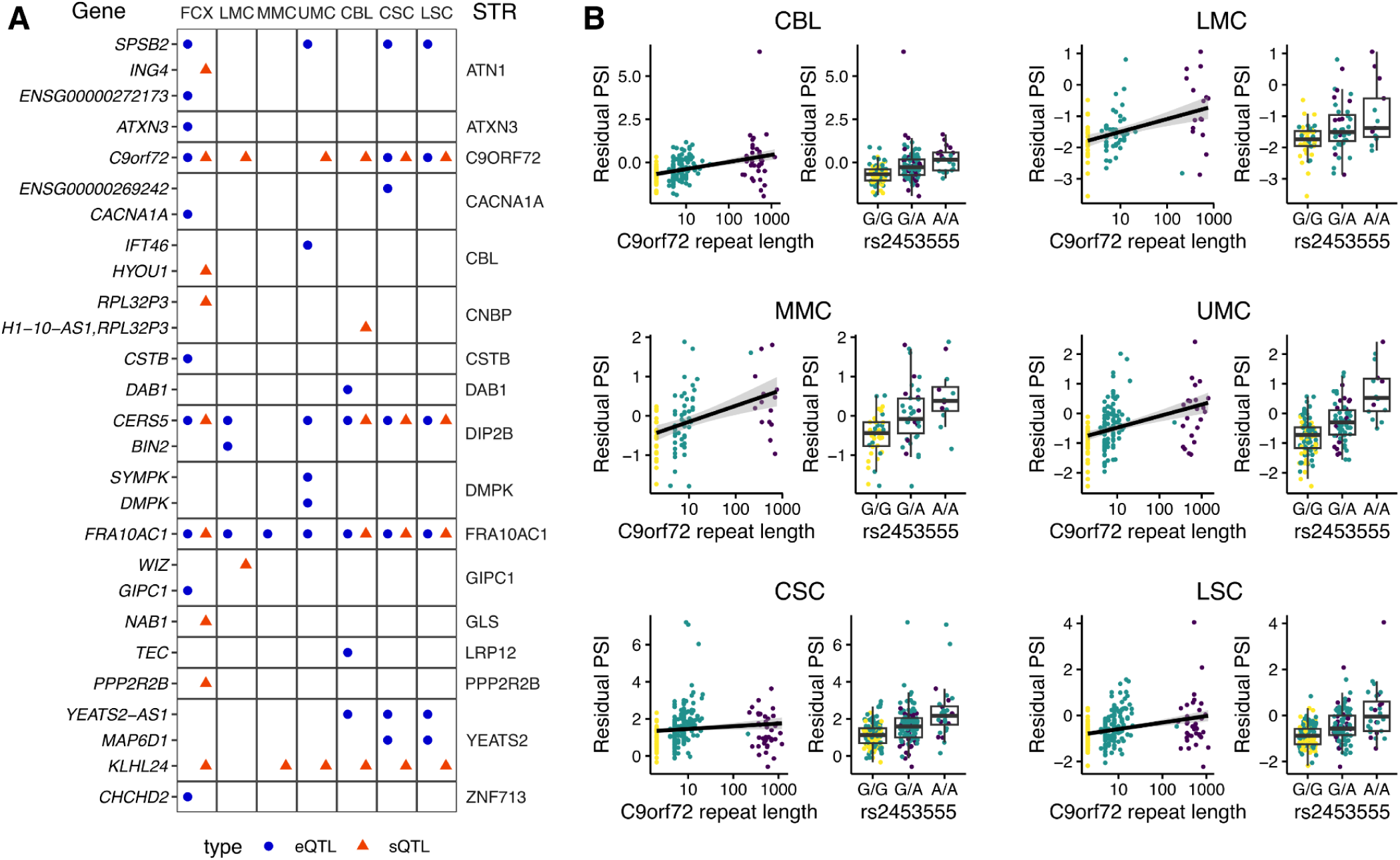
| STR-QTLs. **A**) Full STR-QTL results for all STR-gene pairs at qvalue < 0.05. **B**) The *C9orf72* repeat expansion and lead GWAS SNP associate with increased exon 1a - exon 2 splicing across all tissues.

**Figure S13.**
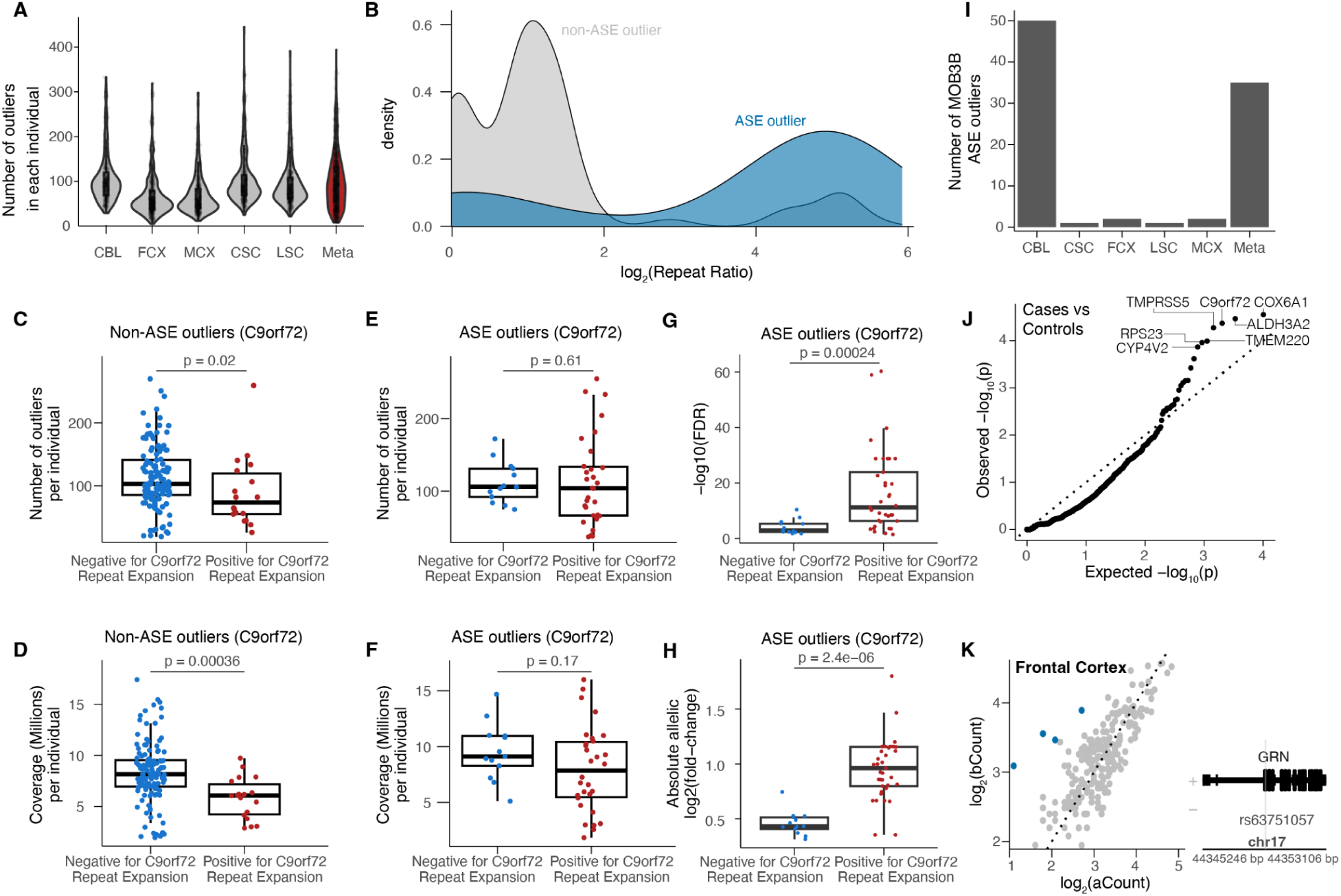
Additional characterization of ASE outliers. **A)** Violin plots showing the distribution of the number of ASE outliers per individual in each of the 5 tissues and the meta-analysis (shown in red). **B)** Density plots showing the distribution of the log_2_(repeat ratio) for *C9orf72* ASE outliers (blue) vs *C9orf72* nonOutliers (gray). **C)** Boxplots showing the distribution of the number of ASE outliers in non-ASE outliers for *C9orf72* that are negative or positive for the repeat expansion. P-value calculated from a Wilcoxon Rank Sum test. **D)** Boxplots showing the distribution of the genome-wide coverage at heterozygous sites in non-ASE outliers for *C9orf72* that are negative or positive for the repeat expansion. P-value calculated from a Wilcoxon Rank Sum test. **E)** Boxplots showing the distribution of the number of ASE outliers in *C9orf72* ASE outliers that are negative or positive for the repeat expansion. P-value calculated from a Wilcoxon Rank Sum test. **F)** Boxplots showing the distribution of the genome-wide coverage at heterozygous sites in *C9orf72* ASE outliers that are negative or positive for the repeat expansion. P-value calculated from a Wilcoxon Rank Sum test. **G)** Boxplots showing the adjusted p-values from the meta-analysis for C9orf72 in C9orf72 ASE outliers that are either negative or positive for the repeat expansion. **H)** Boxplots showing the absolute aFC from the meta-analysis for C9orf72 in C9orf72 ASE outliers that are either negative or positive for the repeat expansion. **I)** Barplots showing the number of individuals that are ASE outliers for *MOB3B* across various tissues. **J)** Quantile-quantile plot showing genes enriched for being ASE outliers in ALS cases vs controls. P-values were calculated from one-sided Fisher’s exact test. **K)** Scatter plot showing the log_2_(haplotype A) vs log_2_(haplotype B) counts for *GRN*, with individuals significant for ASE shown in blue (left). Gene tracks showing the location of frameshift mutation rs63751057 (right) + and - represent the plus and minus strands respectively.

